# TUCAN: Ultra-fast methylation-based classification of pediatric solid tumors and lymphomas

**DOI:** 10.64898/2026.03.24.26348466

**Authors:** Merel Jongmans, Marc van Tuil, Esmée de Ruijter, Laura Hiemcke-Jiwa, Uta Flucke, Ronald de Krijger, Marijn Scheijde-Vermeulen, Pascal Kusters, Roelof van Ewijk, Hans Merks, Max van Noesel, Marc Pagès-Gallego, Carlo Vermeulen, Bastiaan Tops, Jeroen de Ridder, Lennart Kester

## Abstract

The high heterogeneity of pediatric cancers presents significant diagnostic challenges, underscoring the need for accurate classification. Although molecular profiling supports first-line diagnostics and guides treatment, it can delay final diagnosis. While Nanopore-based methylation analysis has enabled rapid CNS tumor diagnosis, its application to pediatric solid tumors and lymphomas has remained largely unexplored. We developed Tucan, a deep-learning classifier trained on 3,818 methylation array profiles representing 84 subtypes, designed to classify tumors from sparse Nanopore methylation data. In retrospective validation (n=514), Tucan generated confident predictions (CFT ***≥*** 0.7) within 30 minutes of sequencing in 385 cases, achieving 372 correct diagnoses (F1-score: 0.98). In prospective testing (n=74; 63 classifiable), 52 samples reached the confidence threshold with 96% accuracy, confirming the original diagnosis in 47 cases and correctly refining or revising it in three. Together, Tucan enables rapid, high-confidence molecular classification of pediatric solid tumors and lymphomas.

## 1 Introduction

Solid tumors, including the lymphomas, account for approximately 40% of all pediatric malignancies and encompass a wide range of distinct subtypes, contributing to significant biological and clinical heterogeneity(Steliarova-Foucher et al, 2017; HESHAM et al, 2014; Pfister et al, 2022). An accurate and timely diagnosis of these heterogeneous tumortypes is essential to determine the appropriate treatment. Despite the high prevalence of solid tumors and lymphomas in children with cancer, pediatric cancer itself is relatively rare (Radhi et al, 2015; Mullen et al, 2021). The combination of high heterogeneity with relatively small sample sizes makes accurate diagnosis of these tumor types particularly challenging.

Figure 1 illustrates the typical diagnostic timeline for pediatric patients, in which diagnosis is based on multiple complementary layers of information. Following biopsy, tissue is processed for histological evaluation, including a Hematoxylin & Eosin stain (H&E) and immunohistochemistry (IHC), which guides the initial differential diagnosis. However, the lack of well-defined histopathological characteristics and the large number of rare tumor entities induces high interobserver variability and increases risk of misclassification (Magro et al, 2015). To refine these preliminary diagnoses, targeted molecular assays (e.g., FISH, PCR) are commonly used to confirm or exclude specific diagnoses (Figure 1) (Pei et al, 2023). When these targeted approaches are inconclusive, more comprehensive profiling techniques such as next-generation sequencing (NGS) or methylation arrays are employed to resolve diagnostically challenging cases (Figure 1) (Malone et al, 2020; Pei et al, 2023). These methods offer additional insights into tumor-specific genetic and epigenetic signatures, enabling more precise classification and reducing diagnostic interobserver ambiguity (Malone et al, 2020; Satam et al, 2023). For example, undifferentiated small round cell sarcomas may appear morphologically similar yet are defined by distinct molecular alterations, such as EWSR1–FLI1 fusions, BCOR alterations, or CIC–DUX4 fusions, each representing a separate WHO entity (WHO Classification of Tumours Editorial Board, 2020; Antonescu et al, 2017; Borislav A. Alexiev, 2020; Church et al, 2018; WHO Classification of Tumours Editorial Board, 2020; Dehner et al, 2024). In addition to classification, molecular profiling provides critical information relevant for risk stratification (Nguyen et al, 2025). For instance, detection of 1p and 16q loss in Wilms tumor and 1p loss and MYCN amplification in Neuroblastoma is required for therapy stratification (Grundy et al, 1994; Huang and Weiss, 2013), as these CNVs are associated with poor prognosis and guide therapy intensity (Grundy et al, 1994; Huang and Weiss, 2013). Thus, molecular profiling is essential not only for accurate diagnosis but also for therapeutic decision-making (Malone et al, 2020).

**Fig. 1:**
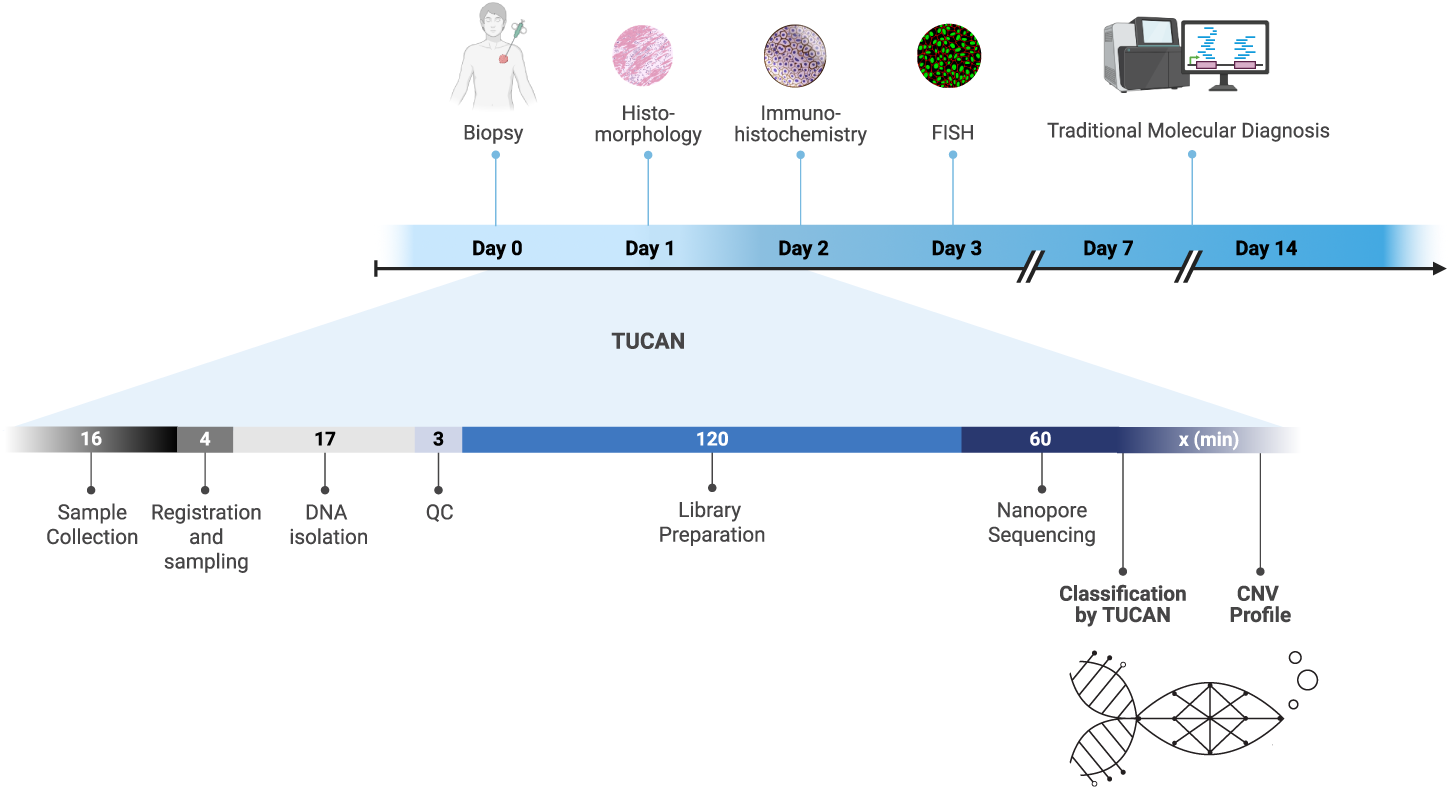
Diagnostic workflow schematic. *Top:* Standard-of-care diagnostic workflow from biopsy to final diagnosis, including sequential histopathological, immunohistochemical, and molecular testing steps. *Bottom:* Diagnostic timeline incorporating Tucan into routine care, demonstrating how rapid Nanopore sequencing accelerates molecular diagnosis and shortens overall turnaround time.

However, molecular profiling can take up to two weeks, potentially delaying treatment initiation and permitting continued tumor progression, thereby increasing exposure to associated risks (Figure 1) (Bhatt et al, 2023). In high-grade pediatric solid tumors, such delays may result in acute, potentially irreversible complications. For instance, tumors located adjacent to or within the spinal canal may cause spinal cord compression with permanent neurological damage, whereas those arising in the head and neck region may lead to vision loss, cranial nerve palsies, or intracranial extension if therapy is not initiated promptly (Śanchez et al, 2024; Gopalakrishnan et al, 2012; FAWZY et al, 2015; Graef et al, 2024; Hayes et al, 2019; Masoudi et al, 2020). These risks highlight the clinical trade-off between achieving precise molecular diagnosis and initiating treatment within a time frame that meaningfully limits disease progression, metastasis, and severe complications.

*Vermeulen et al.* demonstrated how to mitigate this trade-off for the classification of (pediatric) CNS tumors by using methylation profiles acquired with Nanopore sequencing (Vermeulen et al, 2023). However, same-day molecular classification of pediatric solid tumors and lymphomas has not yet been addressed. Here, we developed Tucan (solid Tumor Classification using Nanopore), a deep-learning classifier that integrates rapid Nanopore-based methylation classification and CNV detection directly into the standard-of-care diagnostic workflow and which enables turn-around-times of under 24 hours for molecular classification (Figure 1). We demonstrate the use of Tucan as a powerful and easily implementable diagnostic tool, designed to aid diagnostic decision-making through high-confidence tumor classification (Figure 1). Tucan has been trained on a comprehensive cohort of 3,818 methylation array profiles, covering 84 WHO-defined tumor entities, representing the molecular heterogeneity of pediatric solid tumors and lymphomas. We validated Tucan on 514 retrospective and 74 prospective Nanopore samples, demonstrating Tucan’s classification performance and showcasing its potential clinical utility.

## 2 Results

### 2.1 Comprehensive Data Curation Yields a Cohort Representative of the Diversity of Pediatric Solid tumors and Lymphomas

For the development of Tucan, we curated a large and diverse reference cohort of pediatric solid tumors and lymphomas, defined according to the WHO classification of tumors. Due to the limited availability of Nanopore-based methylation data, the model was trained on methylation array data adapted to mimic Nanopore-derived methylation profiles. The dataset comprised 3,818 samples derived from 25 sources, including GEO, TCGA, TARGET, and the Princess Máxima Center Biobank, profiled using 450K, EPIC, or EPICv2 arrays (Supplementary Table S2). In total, the cohort covered 84 tumor subtypes and six control groups across 21 tumor families (Fig. 2). At the subtype level, several entities were removed or merged based on their methylation characteristics. Specifically, undifferentiated sarcoma, rhabdomyosarcoma-like sarcoma, and Langerhans cell histiocytosis were excluded, while neuroblastoma and ganglioneuroblastoma, as well as BCOR sarcoma and clear cell sarcoma of the kidney, were merged into unified classes based on their highly similar methylation profiles (Supplementary Fig. S1). To ensure label reliability, an iterative neighborhood-based filtering strategy was applied, removing samples with low concordance among nearest neighbors (Methods 4.2). This process led to the exclusion of three subtypes that failed to form distinct methylation clusters or showed poor consistency between pediatric and reference samples (inflammatory myofibroblastic tumor (IMT), translocation associated renal cell carcinoma (tRCC), and perivascular epithelioid cell tumor (PEComa)) (Supplementary Fig. S1). The final cohort captures the biological diversity of pediatric solid tumors and lymphomas, while preserving robust subtype-specific methylation patterns. Sample counts per subtype ranged from 5 to 354, reflecting real-world disease prevalence and inherent class imbalance (Fig. 2 and Supplementary Table S2).

**Fig. 2:**
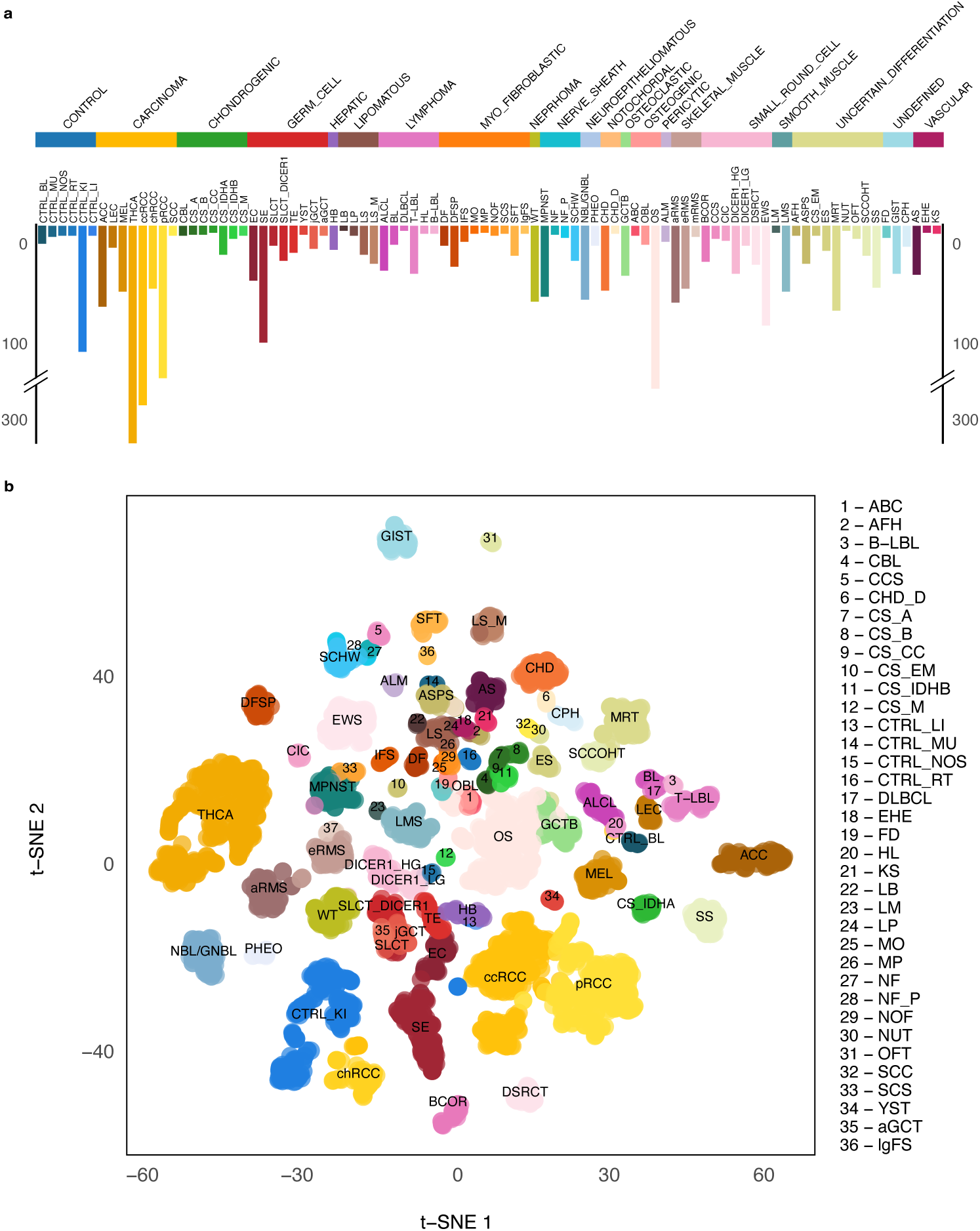
Cohort Overview - Methylation Array. **(a)** Overview of the methylation array training data. *Top:* Bar plot showing the distribution of families, where the width of each bar represents the number of distinct subtypes within that family. *Bottom:* Histogram displaying the number of training samples per subtype. **(b)** t-SNE visualization of the methylation array training data based on the 5000 most variable CpG sites. Samples are colored according to their subtype and labeled with either an abbreviation or a numeric code. The legend on the right provides the mapping of the numeric codes to their respective subtype abbreviations.

### 2.2 Validation fold was used for Model Calibration and Threshold Selection

To meet diagnostic turnaround requirements, there is only time for Nanopore sequencing runs that yield sparse CpG coverage. To account for this, we applied a simulation strategy adapted from *Vermeulen et al.*, which is aimed at delivering accurate classification based on sparse data. This approach was applied to the methylation microarray cohort and the resulting simulated data was used to train Tucan using four-fold cross-validation. Four independent models were generated by rotating training, calibration, and evaluation folds, maximizing data usage while maintaining robust performance estimates. Validation folds were used to assess model calibration and to define confidence thresholds. The models were trained on simulated datasets representing 10,000–39,500 CpG sites, with classification performance increasing with coverage (Supplementary Fig. S2). Calibration was assessed on the validation folds using the expected calibration error (ECE), yielding a value of 0.0093, indicating close agreement between expected and observed accuracy (Supplementary Fig. S2). This low ECE was primarily driven by a strong concentration of predictions in the highest confidence bin, where observed accuracy closely matched expected confidence (Supplementary Fig. S2). Lower confidence bins contained relatively few samples and showed no evidence of systematic miscalibration (Supplementary Fig. S2). Given the well-calibrated confidence estimates and the dominance of high-confidence predictions, no additional calibration was applied. A confidence threshold (CFT) of 0.95 was selected for subsequent analyses to ensure high reliability of the classifications.

### 2.3 Simulation-Based Training Enables Accurate Molecular Classification from Sparse Methylation Array Data

Tucan was evaluated on four independent hold-out test sets using simulated sparse methylation profiles comprising 10,000 CpG sites. At this coverage level, Tucan achieved a mean F1 score of 0.98, demonstrating robust predictive performance, with misclassifications appearing mainly between related entities (Fig. 3). Although overall performance was high, a small number of subtypes remained more challenging to classify. These included chondrosarcoma IDH group B (CS B), lipoblastoma (LB), diffuse large B-cell lymphoma (DLBCL), and kinase driven spindle cell sarcoma (SCS), which all have limited representation in the training data and close epigenetic similarity to more prevalent subtypes within their families (*N_CS B_* = 9, *N_LB_* = 5, *N_DLBCL_* = 5, *N_SCS_* = 8) (Fig. 2 and 3). Together, these results show that, using a CFT *≥* 0.95, Tucan enables accurate classification from sparse array-derived methylation profiles.

**Fig. 3:**
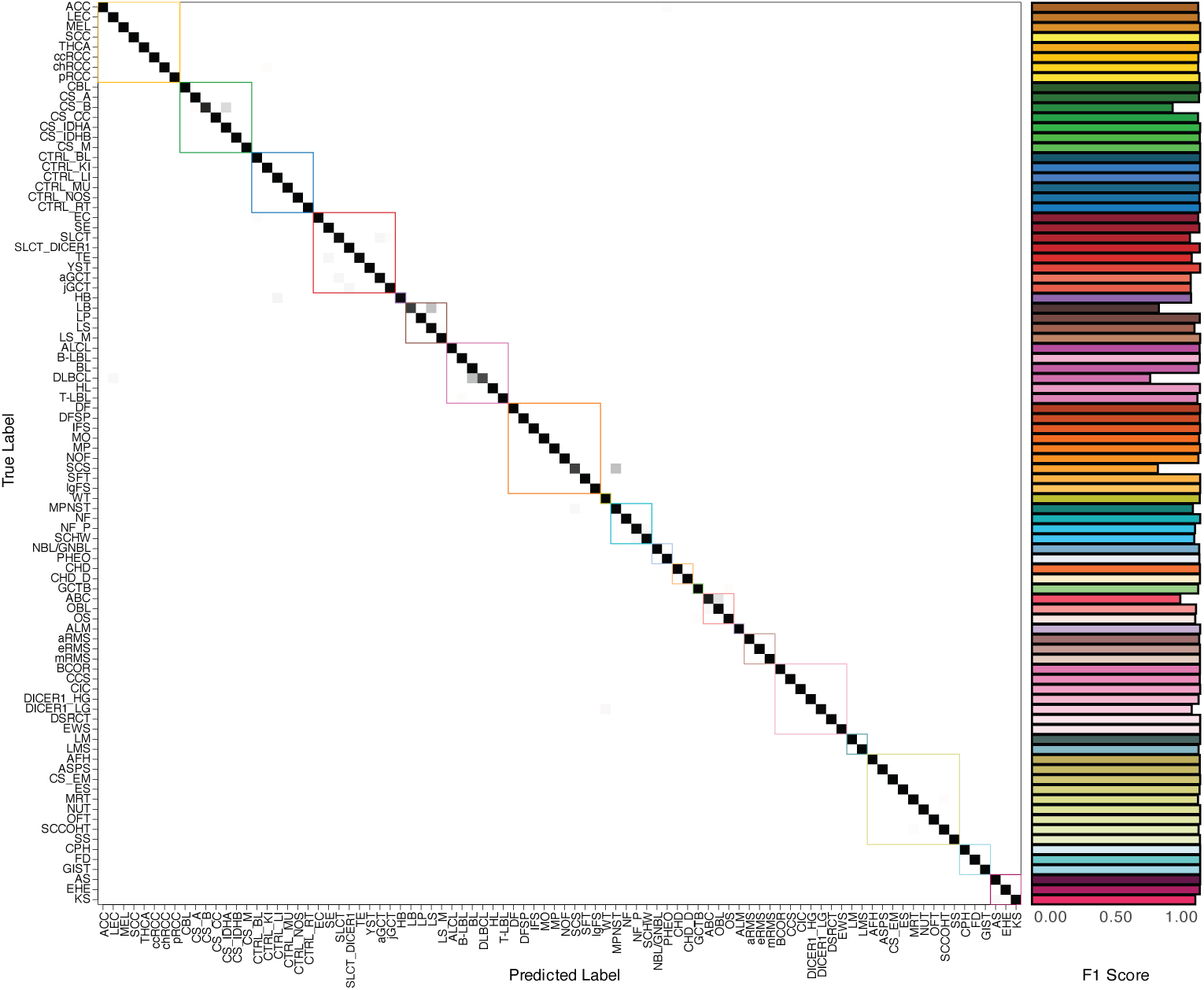
Performance of Tucan - Methylation Array Test Data. Correlation matrix of the methylation array test data, evaluated at a sequencing depth of 10,000 CpG sites and a confidence score of *≥* 0.95. Subtypes are color-coded, with true labels displayed along the vertical axis and predicted labels along the horizontal axis. The corresponding F1-scores for each subtype are displayed to the right of the matrix.

### 2.4 Retrospective Validation Confirms Tucan’s Accuracy and Outperforms the Heidelberg Classifier

To further assess Tucan’s performance on independent methylation array data, we retrospectively validated the model on 114 additional cases and compared its performance to the Heidelberg Sarcoma Classifier (Koelsche et al, 2021). Because the Heidelberg classifier is restricted to sarcoma entities and Tucan does not include IMT samples, the comparative analysis was limited to 95 classifiable cases. For this evaluation, Tucan was applied using a CFT *≥* 0.95, consistent with the threshold established during array-based validation. Similarly, the Heidelberg classifier was applied using its recommended CFT *≥* 0.9 (Koelsche et al, 2021). On confidently classified samples (*N_T ucan_* = 71 and *N_Heidelberg_* = 66) Tucan achieved an accuracy of 100%, compared to 93% for the Heidelberg classifier (Supplementary Fig. S11). Taken together, these results show that Tucan achieves high accuracy on independent methylation array data and outperforms the current gold standard in terms of the number of classifiable samples and performance.

### 2.5 Tucan Enables Ultra-Fast and Accurate Molecular Classification on Nanopore Sequencing Data

Tucan was retrospectively validated on nanopore sequencing data from 514 samples representing 49 tumor subtypes. Samples were generated on PromethION or MinION flow cells, reaching 10,000 CpG sites within approximately 10 and 30 minutes, respectively, when sequencing a single sample per flow cell. To account for stochastic variation introduced by sparse methylation coverage, each of the four Tucan submodels was run 100 times on independent random subsets of 10,000 CpG sites, and the final classification was based on the mean prediction scores across all iterations and submodels, yielding a single consensus classification per sample.

Applying the CFT of 0.95 established during array-based validation, 54% (275/514) of samples yielded high-confidence predictions, with a conditional accuracy of 99%. Notably, most predictions below this threshold were still correct, indicating that the model’s reported confidence scores tended to underestimate classification accuracy on Nanopore data. (Fig. 4 and Supplementary Fig. S4). Lowering the confidence threshold to 0.8 or 0.7 substantially increased recall, while conditional accuracy remained high (*Acc_CF T_* _=0.7_ = 96.62% and *Acc_CF T_* _=0.8_ = 97.7%), reflecting a favorable trade-off between recall and performance (Fig. 4 and Supplementary Fig. S4). This behavior is consistent with a modest domain shift between the simulated array-derived training data and Nanopore-derived methylation profiles. In the array-based setting, predictions were predominantly assigned to the highest confidence bin, resulting in close agreement between recall and conditional performance across thresholds (Supplementary Fig. S2). In contrast, Nanopore-based predictions were more broadly distributed across confidence bins, with slight underconfidence at the upper end of the confidence scale (Supplementary Fig. S3). Importantly, predictions falling just below the original CFT (in the 0.7–0.8 and 0.8–0.9 ranges) retained high conditional accuracy, indicating preserved discriminative performance despite shifted confidence estimates (Supplementary Fig. S3). Accordingly, while a confidence threshold of 0.95 is appropriate for array-based data, Nanopore-derived classifications benefit from a lower operational threshold. In routine practice, a threshold of 0.7 provides a favorable balance between recall and accuracy, reflecting preserved discriminative performance despite modest underconfidence. Misclassifications observed at these CFTs were largely restricted to biologically related tumor entities, most frequently within the lymphoma and germ cell tumor families, consistent with known overlap in their methylation profiles and, in some cases, limited training representation (*N_DLBCL_* = 5, *N_HL_* = 8, *N_NF_* = 9) (Fig. 2 and 4). Although the retrospective cohort does not capture the full spectrum of rare pediatric tumors, it is representative of the tumor distribution encountered in routine clinical diagnostics. Together, these results demonstrate that Tucan enables rapid and accurate molecular classification from sparse Nanopore methylation data.

**Fig. 4:**
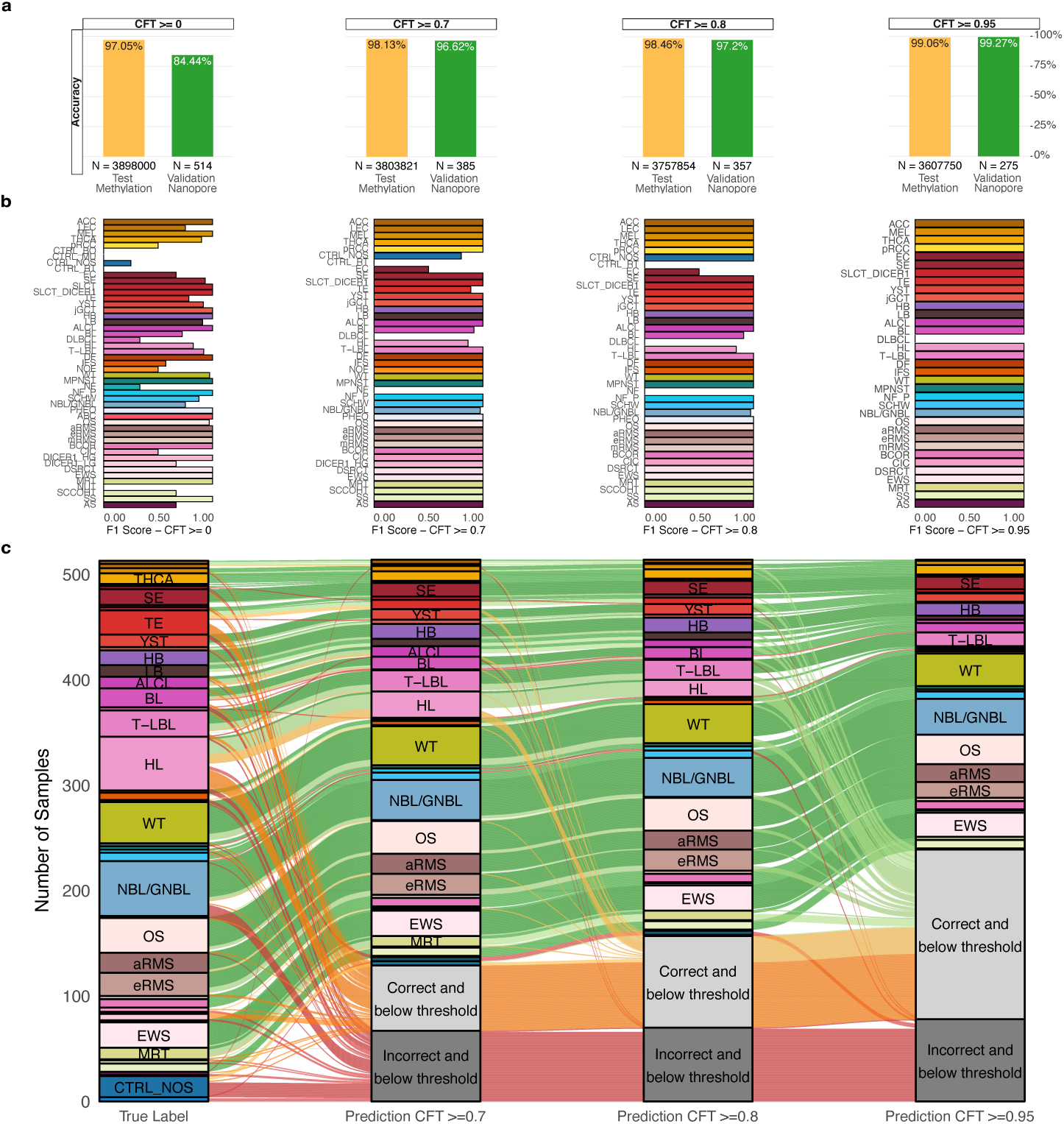
Performance of Tucan - Retrospective Nanopore Samples. **(a)** Accuracy on the simulated methylation array test data (yellow) and retrospective Nanopore samples (green) at a sequencing depth of 10,000 CpG sites across varying confidence thresholds. The plot illustrates the trade-off between decreasing global recall and increasing conditional performance. **(b)** F1-scores by subtype for retrospective Nanopore samples across varying confidence thresholds, showing improved performance at higher thresholds. Bars are colored by subtype. **(c)** Alluvial plot showing Tucan’s predictions at confidence thresholds (CFTs) of 0.7, 0.8, and 0.95. The left column represents the true tumor labels, and the three right columns display Tucan predictions at each CFT, colored by subtype. Predictions below the respective CFT are shown in grey. Flows are colored by correctness: green, correct (CFT *≥* 0.8); yellow, correct (CFT: 0.7 – 0.8); orange, correct (CFT *<* 0.7); and red, incorrect.

### 2.6 Shallow Nanopore Sequencing Enables Detection of Prognostically and Diagnostically Relevant CNVs

Copy number variation (CNV) analysis can provide critical information for tumor classification, risk stratification, and therapeutic decision-making in selected entities (Pös et al, 2021; Hahn et al, 2025). We therefore assessed whether shallow Nanopore sequencing could enable rapid and reliable CNV detection to complement the ultra-fast methylation-based classification. CNVs were inferred from read-depth profiles which were normalized using a panel of normals, improving robustness compared to single-sample normalization approaches (Hahn et al, 2025; Ahmad et al, 2023; Quenez et al, 2021) (Supplementary Fig. S5).

Nanopore-derived CNV profiles were compared to whole-exome sequencing (WES) derived CNV profiles. Concordance increased with sequencing depth and plateaued at approximately 100,000 reads, reaching 99% agreement with WES-based CNV calls (Supplementary Fig. S7). Using 2 Mb bins and a copy-ratio threshold of *±*0.3, CNVs were reproducibly detected well above random expectation (Supplementary Fig. S7). While shallow coverage inherently limits sensitivity for small alterations, focal events such as MYCN amplifications were detectable from as few as 10,000 reads, with stable genome-wide CNV profiles achieved at *±*100,000 reads (approximately 45 minutes of sequencing and corresponding to *±*0.06x genome coverage), which we define as the recommended minimum depth (Supplementary Fig. S7).

Although many pediatric solid tumors and lymphomas exhibit few or no CNVs (Supplementary Fig. S6), several clinically relevant subtypes are characterized by distinctive CNV patterns critical for diagnosis and risk stratification (Segers et al, 2013; Huang and Weiss, 2013; Meijer et al, 2024; Smith et al, 2013). For example, the combined loss of 1p and 16q with gain of 1q, associated with poor prognosis in Wilms tumor, was clearly detected, as was the MYCN amplification defining high-risk neuroblastoma (Segers et al, 2013; Huang and Weiss, 2013) (Fig. 5). Beyond risk stratification, CNV profiles can also contribute to diagnosis. Osteosarcomas displayed highly complex, genome-wide copy number alterations that were clearly visible in the Nanopore-derived CNV profiles (Meijer et al, 2024) (Fig. 5). Similarly, the CNV profiles can help distinguish alveolar- and embryonal rhabdomyosarcoma, which differ markedly in genomic instability (Smith et al, 2013) (Fig. 5).

**Fig. 5:**
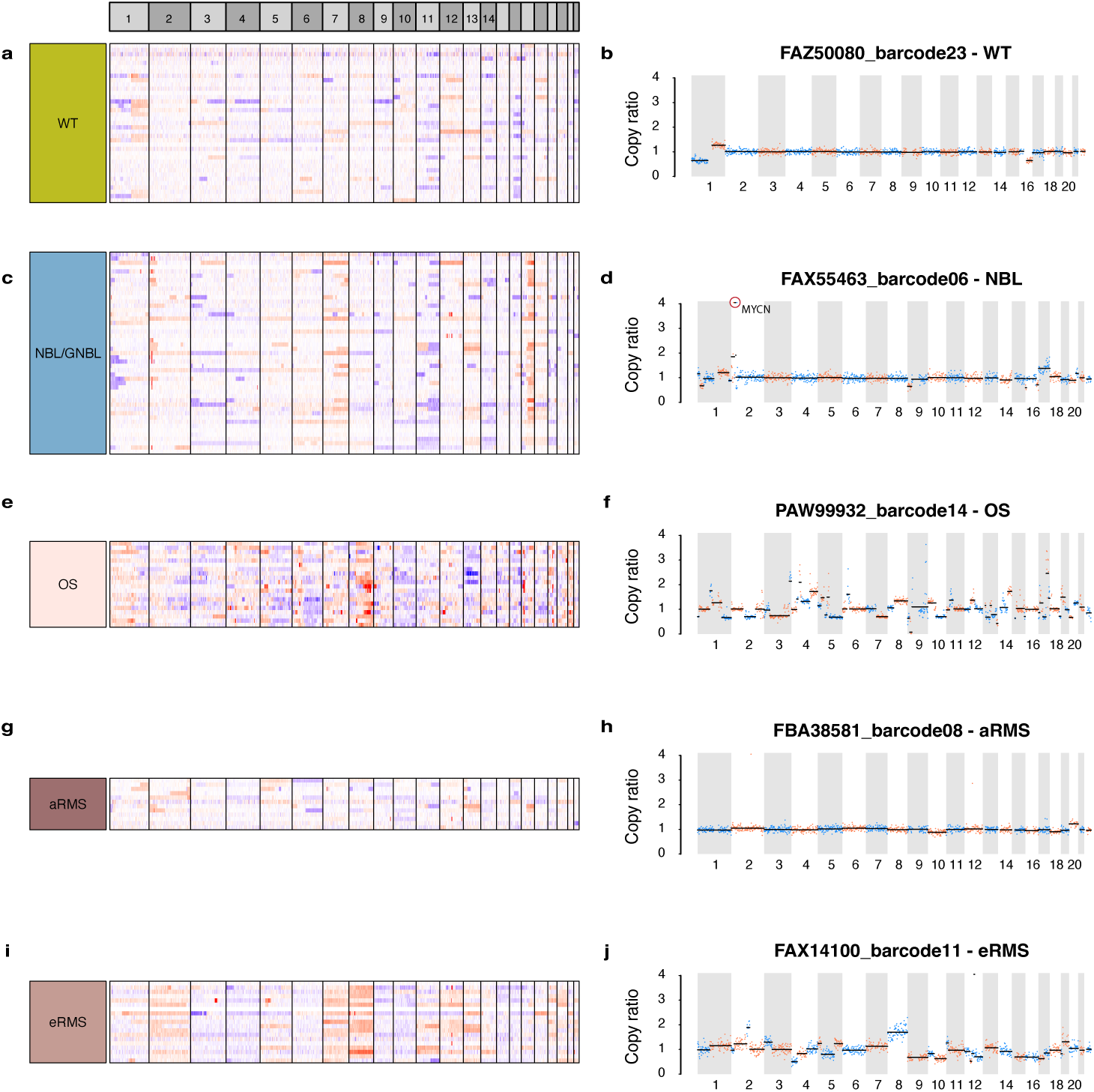
Copy Number Variations - Retrospective Cohort(a, c, e, g, **i)** Heatmaps of copy number variations (CNVs) per chromosome per sample for Wilms tumor (WT), Neuroblastoma/Ganglioneuroblastoma (NBL/GNBL), Osteosarcoma (OS), alveolar rhabdomyosarcoma (aRMS), and embryonal rhabdomyosarcoma (eRMS). Each row represents an individual sample, color-coded by subtype. Columns correspond to chromosomes, with alternating light and dark grey shading for visual clarity. Red indicates copy number gains, blue indicates copy number losses, and white denotes neutral regions. **(b, d, f, h, j)** Representative CNV profiles for the corresponding subtypes. The horizontal axis denotes chromosomes (alternating grey and white blocks), and the vertical axis shows the copy ratio. Dots indicate copy ratio values calculated for 2Mbp bins, with black horizontal lines marking the segmentation boundaries. Dots alternate in blue and red per called segmentation for visual clarity. Subtype-specific examples include: **(b)** WT with poor prognosis, characterized by loss of 1p and 16q and gain of 1q; **(d)** NBL with poor prognosis, characterized by MYCN amplification on chromosome 2; **(f)** OS exhibiting a characteristic CNV pattern with extensive gains and losses; **(h)** aRMS displaying a stable CNV profile; **(j)** eRMS with a CNV profile characterized by whole-chromosome gains and losses.

Together, these results demonstrate that shallow Nanopore sequencing enables rapid and accurate detection of diagnostically and prognostically relevant CNVs, despite limited sequencing depth.

### 2.7 Prospective Validation demonstrates the Clinical Utility of Tucan in Routine Diagnostics

To evaluate clinical performance, Tucan was prospectively validated within the standard diagnostic workflow for 74 patients treated in the Princess Máxima Center between December 2024 and July 2025 (Fig. 1). Biopsy material obtained for routine histopathological and molecular analyzes was also used for Nanopore sequencing. For urgent cases, the time from sample acquisition to start of sequencing was under three hours. Acquisition of the recommended coverage of 10,000 CpG sites, sufficient for high-confidence classification, required on average 15 minutes of sequencing, with an additional 30 minutes to generate a CNV profile. Of the 74 samples analyzed, eleven corresponded to tumor entities not represented in Tucan’s reference set, yielding 63 cases eligible for classification (Supplementary Fig. S8 and S9). Among these classifiable cases, 52 (83%) reached the predefined confidence threshold of 0.7, of which 50 (96%) were correctly classified (Supplementary Fig. S8).

For the eleven samples representing entities absent from the reference set, only one sample exceeded the CFT (*CFT ≥* 0.7) at 10,000 CpG sites (Supplementary Fig. S9). This sample, PMC70, a Langerhans cell histiocytosis, was predicted as control blood. Increasing coverage to 20,000 CpG sites resulted in one additional prediction above the CFT (PMC68 predicted as control NOS) (Supplementary Fig. S9). Importantly, as both samples were classified as control, these misclassifications have no clinical impact. Further increasing sequencing depth led to additional high-confidence misclassifications despite the absence of the correct entity in the reference set, indicating that increasing CpG coverage beyond the recommended threshold may promote overconfident misclassifications when no matching class is available (Supplementary Fig. S9). Importantly, at the recommended coverage of 10,000 CpG sites, all except one of these samples did not reach the CFT, appropriately reflecting the absence of a matching class in the reference model (Supplementary Fig. S9).

Among the 63 eligible cases, there were two misclassifications above CFT, including a yolk sac tumor, misclassified as hepatoblastoma (PMC61), and an embryonal rhabdomyosarcoma misclassified as alveolar rhabdomyosarcoma (PMC26) (Supplementary Fig. S8). Notably, the Nanopore-derived CNV profile of the latter case showed a CNV profile characteristic of embryonal rhabdomyosarcoma, indicating concordance at the copy-number level (Supplementary Fig. S10). Of the 50 correctly classified cases, 47 were concordant with the initial pathological diagnosis, while in three cases the initial diagnosis was refined or reclassified following Tucan’s prediction. Four of the clinical cases will be highlighted below, demonstrating how Tucan provided molecular insights that altered or refined routine diagnostic decision-making.

#### 2.7.1 Case 1 - Reclassification to BCOR sarcoma

A girl aged 11–15 years presented with a lumbar soft-tissue mass without further symptoms. MRI revealed a 7.5 × 5 × 6 cm lesion in the erector spinae at the L5/S1 level, with destruction of the L5 spinous process and extension into the spinal canal toward the S1 neuroforamen. Staging with chest CT and [^^^18F]FDG-PET showed no metastatic disease. Histopathological examination demonstrated a monotonous proliferation of primitive spindle cells within a myxoid stroma (Fig. 6). Based on morphology and immunohistochemistry, the differential diagnosis included an NTRK-rearranged spindle cell neoplasm (due to the spindle-like morphology and NTRK IHC positivity), malignant peripheral nerve sheath tumor, or synovial sarcoma (Fig. 6). Tucan classified the sample as a BCOR sarcoma, which in retrospect aligned with the observed NTRK immunoreactivity, likely reflecting upregulation of NTRK3 mRNA rather than a NTRK fusion (Fig. 6). Detection of a BCOR fusion by RNA sequencing 2 weeks later confirmed Tucan’s classification, establishing this rare diagnosis that had not been considered in the initial differential diagnosis, after which therapy was initiated.

**Fig. 6:**
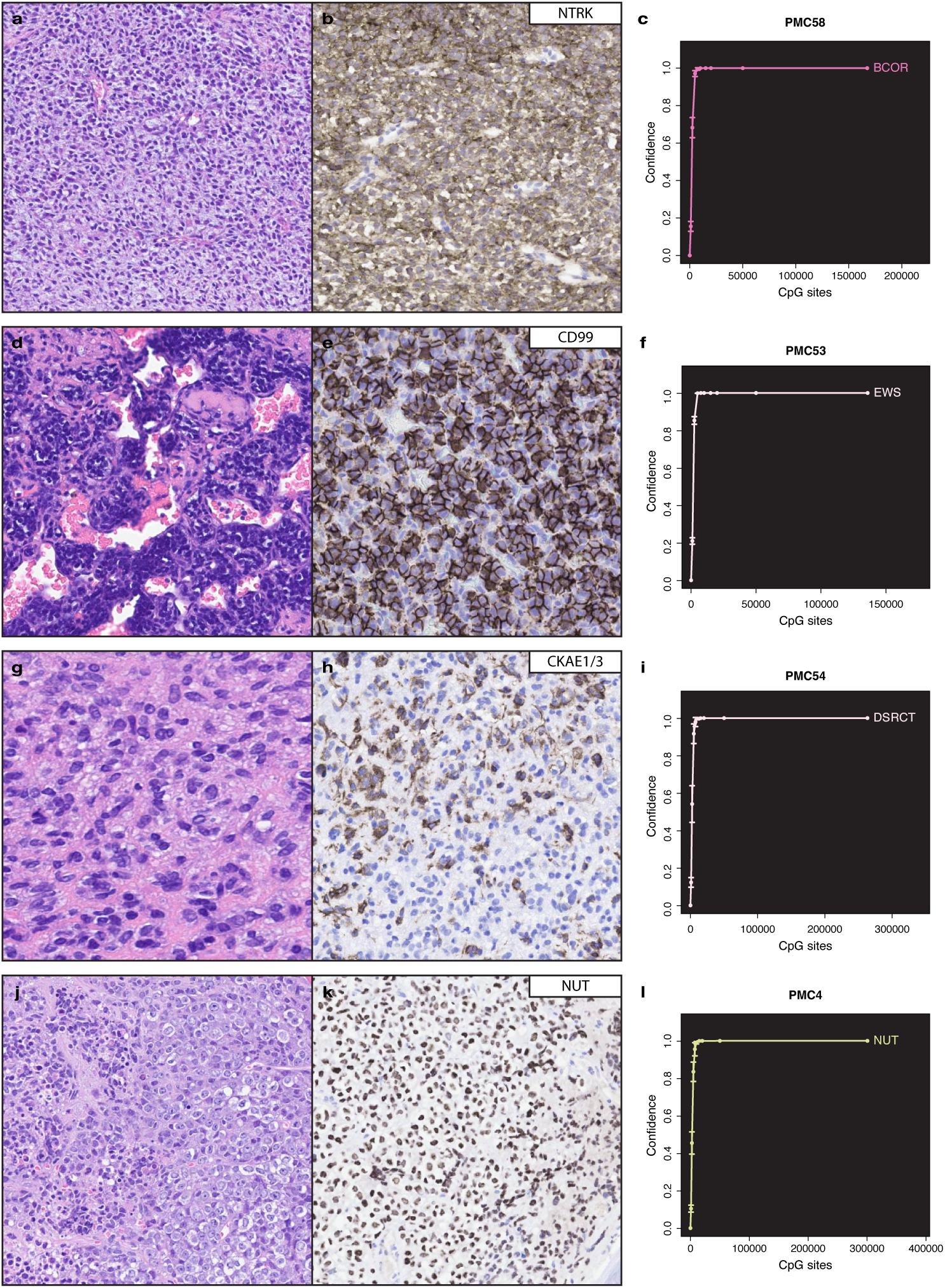
Diagnostic test results for four individual cases. (a, b, **c)** PMC58 - BCOR sarcoma (BCOR) **(d, e, f)** PMC53 - Ewing Sarcoma (EWS) **(g, h, i)** PMC54 - Desmoplastic small round cell sarcoma (DSRCT) **(j, k, l)** PMC4 - NUT Carcinoma (NUT) **(a, d, g, j)** Hematoxylin and eosin (H&E) staining: the first line of diagnostic testing and guiding subsequent analyses. **(b, e, h, k)** Immunohistochemical stainings: **(b)** NTRK, **(e)** CD99, **(h)** CKAE1/3, **(i)** NUT. **(c, f, i, l)** Classification by Tucan. The horizontal axis shows the number of CpG sites covered, and the vertical axis the corresponding confidence score. Each dot with error bars represents the measured confidence score at a given sequencing depth for the given classification. Classifications are colored by subtype.

#### 2.7.2 Case 2 - Diagnostic refinement to EWS

A girl aged 16–20 years presented with a painful, enlarging mass of the upper leg. MRI revealed a large, heterogeneous soft-tissue lesion on the anterolateral side of the right thigh, with focal cortical involvement of the femur, raising suspicion of a bone sarcoma. Staging identified nodal involvement and a solitary bone metastasis. Histopathological evaluation showed a multinodular proliferation of primitive tumor cells with hemangiopericytoma-like vessels and foci of chondroid matrix (Fig. 6). CD99 demonstrated diffuse, strong membranous staining. Differential diagnosis included mesenchymal chondrosarcoma (due to chondroid matrix and hemangiopericytoma-like vasculature), Ewing sarcoma (due to primitive morphology and CD99 positivity), and undifferentiated small round cell sarcoma (Fig. 6). Tucan classified the tumor as Ewing sarcoma (EWS), a result that was confirmed later by RNA sequencing (Fig. 6).

#### 2.7.3 Case 3 - Reclassification to DSRCT

A boy aged 6–10 years presented with abdominal discomfort, nausea, and pain. Ultrasound revealed a left-sided abdominal mass measuring 5.0 × 7.0 × 9.4 cm, located adjacent to the colon but not arising from any specific organ. MRI showed a single solid lesion with mild diffusion restriction, raising suspicion for several malignant as well as borderline or benign tumors. The mass was strongly ^18^F-FDG–avid on PET imaging, without evidence of peritoneal or distant metastases, and chest CT was normal. A core needle biopsy was performed and showed a round cell tumor with spindle type nuclei without prominent nucleoli with eosinophilic cytoplasm and low mitotic activity (Ki-67 *≈* 2%). Immunohistochemistry showed diffuse SMA and desmin positivity and partial CKAE1/3 expression, whereas ALK, CD117, DOG1, Caldesmon, MyoD1, Myogenin, CD99, S100, and CD34 were negative (Fig. 6). SDHA and SDHB expression was retained. Based on morphology and staining, the differential diagnosis included leiomyomatous tumor, myofibroblastic tumor, gastrointestinal stromal tumor (GIST), and GLI-altered tumor, but no conclusive tumortype could be assigned (Fig. 6). Tucan identified the tumor as a Desmoplastic Small Round Cell tumor (DSRCT), which was subsequently confirmed by RNA sequencing, which revealed an EWSR1::WT1 rearrangement (Fig. 6).

#### 2.7.4 Case 4 - Tucan Enables Accurate Diagnosis in a Time-Sensitive Clinical Scenario

A girl aged 11–15 years presented with rapidly progressive dyspnea and clinical signs of superior vena cava syndrome. Imaging revealed a large anterior mediastinal mass with extensive skeletal and pulmonary metastases. Because of impeding respiratory failure, an immediate diagnostic biopsy was not feasible. Empiric chemotherapy with cisplatin, ifosfamide, and doxorubicin was initiated to cover the leading differential diagnoses, which included NUT carcinoma, malignant germ cell tumor, and soft tissue sarcoma. After an initial partial response, the patient’s respiratory condition deteriorated within two weeks due to tumor progression, allowing only a limited trephine biopsy of a pelvic skeletal metastasis. Histopathological evaluation showed a diffuse proliferation of primitive tumor cells with vesicular nuclei, prominent nucleoli, and a destructive growth pattern. Small foci of abrupt squamous differentiation were noted in the frozen section (Fig. 6). Immunohistochemistry demonstrated diffuse granular nuclear NUT positivity, with CKAE1/3 and P40 also diffusely positive, findings pathognomonic for NUT carcinoma (Fig. 6). Nanopore sequencing, analyzed using Tucan, confirmed NUT carcinoma within hours, aligning fully with the histopathological diagnosis (Fig. 6). Given the widely metastatic and rapidly progressive nature of the disease, which was refractory to empiric chemotherapy, the prognosis was considered dismal, and further systemic therapy was not pursued. The patient succumbed to the disease five days after molecular confirmation.

Together, these prospective analyses demonstrate that Tucan enables rapid and accurate molecular classification across a range of routine and time-critical diagnostic scenarios.

## 3 Discussion

Here, we presented Tucan, a rapid machine-learned classifier enabling accurate same-day molecular classification of pediatric solid tumors and lymphomas. Tucan is designed as an easy-to-implement tool for routine diagnostic workflows. By combining methylation-based classification with ultra-rapid Nanopore sequencing, Tucan addresses the clinical trade-off between diagnostic speed and molecular precision, providing robust molecular insights within clinically actionable time frames.

As a foundation for Tucan, we assembled an extensive and carefully curated reference cohort for model development. Through iterative neighborhood-based filtering and subtype consolidation, we prioritized biologically coherent methylation profiles over annotation-based diagnostic labels, excluding entities that failed to form stable clusters or showed insufficient consistency across datasets. Although this reduced the total number of subtypes represented, it strengthened the reliability of the labels and minimized the risk of learning misleading patterns.

Comparison with the Heidelberg Sarcoma Classifier contextualizes Tucan’s performance relative to the current gold standard in methylation-based classification. The Heidelberg classifier relies on array-based data and is limited to sarcoma entities, whereas Tucan is designed to analyze both array- and Nanopore-derived methylation profiles and covers a broader diagnostic spectrum, extending beyond sarcomas to include all pediatric solid tumors and lymphomas. Notably, when evaluated on array-derived methylation data, Tucan matched or exceeded the performance of the Heidelberg classifier on our internal retrospective cohort, achieving 100% accuracy at its predefined confidence threshold. This comparison was, however, limited to an in-house dataset and may therefore reflect local cohort composition and laboratory workflows, underscoring the need for further multi-center validation. Nonetheless, these findings demonstrate that Tucan generalizes well beyond its primary sequencing modality.

Retrospective validation on Nanopore data demonstrated Tucan’s performance on rapidly generated data while simultaneously revealing a domain shift relative to array-based methylation data. This shift primarily manifested as lower model confidence, with Nanopore predictions distributed across lower confidence bins. Importantly, this behavior does not represent a major limitation and is accommodated through adjusted confidence thresholds without compromising clinical reliability. Although the retrospective cohort does not encompass the full spectrum of rare pediatric tumor entities, it closely mirrors the distribution encountered in routine diagnostics, supporting the relevance of these findings. Consistent with known biological overlap in methylation landscapes, the most challenging classifications were observed within the lymphoma and germ cell tumor families, particularly for closely related subtypes or entities with limited training representation.

Beyond methylation-based classification, Tucan provides additional diagnostic value through the generation of CNV profiles. Within a few hours, this enables the detection of clinically relevant large-scale genomic alterations with established prognostic and diagnostic value, including the MYCN amplification in neuroblastoma and 1p/16q loss or 1q gain in Wilms tumor, as well as copy-number patterns that aid in distinguishing embryonal from alveolar rhabdomyosarcoma. While this approach is inherently limited to large-scale structural alterations and does not capture single-nucleotide variants or gene fusions, it provides complementary molecular insights that can substantiate or refine the classification results.

The prospective validation of Tucan within our routine diagnostic workflows demonstrates its clinical feasibility and added value. In most cases, Tucan rapidly confirmed the initial pathological diagnosis, thereby reinforcing diagnostic confidence and enabling timely treatment initiation. In a subset of cases, Tucan enabled diagnostic refinement or reclassification, redirecting diagnoses towards clinically more appropriate entities. The parallel availability of CNV profiles provided additional molecular context, either strengthening or redefining diagnoses, particularly in cases with ambiguous histopathology. Several limitations should nevertheless be acknowledged. Although Tucan covers the majority of WHO-defined pediatric solid tumor and lymphoma entities, certain rare subtypes are currently not included, either due to insufficient high-quality training data or because their methylation profiles lack sufficient distinctiveness for reliable classification. As a consequence, these entities cannot yet be reliably classified and may remain unclassified or be at risk of misclassification. Although our prospective validation indicates that non-represented subtypes typically fail to reach the predefined confidence threshold within the recommended sequencing time (10.000-20.000 CpG), excessive sequencing input beyond this range increases the risk of overconfident misclassifications and is therefore highly discouraged. Importantly, despite these constraints, Tucan proved particularly valuable in histopathologically ambiguous and/or urgent, time-critical cases, enabling same-day molecular classification. Its results should, whenever feasible, be interpreted within an integrated diagnostic framework that incorporates histopathology. While we have demonstrated the added value of Tucan in a technologically advanced pediatric oncology center, it’s impact may be even greater in settings that lack infrastructure for comprehensive genomic profiling. In low-and middle-income countries in particular, the relatively low investment requirements and operational costs of Nanopore sequencing enable Tucan to serve as a stand-alone molecular classification tool, potentially broadening access to modern molecular diagnostics in resource-limited settings.

In conclusion, Tucan represents a practical and easy clinically implementable molecular diagnostic tool. By providing rapid, reliable, and interpretable molecular classifications, Tucan can support and refine initial diagnoses, guide clinical decision-making, and substantially shorten diagnostic turnaround times, from weeks to hours, thereby mitigating the trade-off between molecular precision and the urgency of initiating timely treatment.

## 4 Methods

### 4.1 Overview of Modeling Framework

We developed Tucan, a multi-platform methylation classifier extending the Sturgeon framework (Vermeulen et al, 2023). Similar to Sturgeon, Tucan is compatible with both Illumina methylation array and Nanopore sequencing data. Whereas the original Sturgeon model was trained on 450K array data for brain tumor classification, Tucan expands the framework to include EPIC and EPICv2 array and to encompass 84 pediatric solid tumor and lymphoma classes. The overall workflow comprises array harmonization, simulation of sparse Nanopore-like profiles, neural network training, and ensemble-based cross-validation.

### 4.2 Data Preparation and Array Harmonization

Raw methylation array data were collected from public and in-house cohorts generated on the Illumina HumanMethylation450 (450K), EPIC, and EPICv2 platforms (Supplementary Table S2). To enable cross-platform integration, we restricted the data to 353232 CpG sites shared among the 450K, EPIC, and EPICv2 arrays, yielding a harmonized feature matrix used for downstream simulations and model training. However, before performing this cross-platform harmonization, we applied several preprocessing and filtering steps to obtain a high-quality and consistently defined training cohort.

The unfiltered cohort comprised 4212 samples representing 90 diagnostic WHO-entities and six control groups, with an overview provided in Supplementary Table S2. The initial step involved excluding diagnostically ambiguous labels: undifferentiated sarcoma, sarcoma RMS-like, and Langerhans cell histiocytosis. In addition, neuroblastoma and ganglioneuroblastoma were merged to resolve inconsistent annotations across datasets.

Subsequently, an iterative filtering strategy was applied to identify and exclude outlier or low-quality samples based on neighborhood consistency within the methylation space. Dimensionality reduction was first performed using Principal Component Analysis (PCA) on the top 5,000 most variable CpG sites, retaining the first 100 principal components. For each sample, the ten nearest neighbors were identified in PCA space, and label concordance among these neighbors was calculated. Labels were removed based on incrementing concordance and decreasing number of cases per subtype for each ten nearest neighbors.

More specifically, samples were iteratively excluded if they exhibited low concordance (*≤* 0, *≤* 1, or *≤* 2 matches) with the neighboring labels. This filtering was repeated across successive rounds, each time applying a lower minimum number of samples per label (*>* 50, *>* 20, *>* 10, and finally *>* 0). Filtering was progressively refined, and round 12—removal of samples with *≤* 2 concordant neighbors for all labels with *>* 0 cases was selected as the optimal trade-off between cleaning the data set and preserving the representation of rare labels.

### 4.3 Simulation of Sparse Nanopore Profiles

To mimic the variability and sparsity of Nanopore methylation sequencing in the training data, each microarray profile was upsampled by generating thousands of simulations that mimic Nanopore reads. For each simulation, the number of reads (*N*) and average read length (*D*) were sampled from empirical distributions representative of typical MinION sequencing throughput. Reads were then randomly assigned genomic start positions in either forward or reverse orientation, and those extending beyond chromosome boundaries were clipped accordingly. Methylation states for CpG sites covered by each simulated read were inferred from binarized array beta values, designating sites as methylated when *β ≥* 0.6. This procedure yielded an upsampled training data set in which array-derived samples exhibit sparsity, variability, and read-level structure closely resembling those observed in Nanopore methylation sequencing.

### 4.4 Neural Network Architecture and Training Strategy

Tucan employs a three-layer fully connected neural network inspired by the Sturgeon model, but adapted for multi-platform compatibility and expanded applicability across pediatric solid tumors and lymphomas (Vermeulen et al, 2023). The input layer was resized to accommodate the 353232 CpG loci shared across the 450K, EPIC, and EPICv2 microarray platforms. The two subsequent layers comprised 256 and 128 dimensions, respectively, each followed by a sigmoid activation function and a dropout rate of 0.5. The final output layer produces probability distributions across 90 diagnostic classes. Model predictions were normalized by softmax activation. Model training followed a supervised multi-class classification framework. Optimization was performed using the AdamW optimizer with a linear warm-up and cosine decay learning rate schedule, a batch size of 256, and 50 training epochs. Dropout layers were retained between all hidden layers to reduce overfitting.

### 4.5 Cross-Validation and Ensemble Inference

The dataset was split into four folds with preserved class distributions for cross-validation. For each cross-validation run, two folds were used for training, one for validation, and one for testing. Repeating this procedure across all fold combinations yielded four independently trained submodels. Simulation seeds were made mutually exclusive across partitions to prevent data leakage (training: 1,000–1,001,000; validation: 500–999; testing: 0–499). For inference, predictions from the four submodels were ensembled by averaging the per-class confidence scores across folds. The class with the highest aggregated mean confidence score was selected as the final output.

### 4.6 Score calibration

To assess interpretability, we evaluated the calibration of the classification scores, although ultimately no calibration step was applied. The second data fold was reserved exclusively for this evaluation. The calibration performance was examined by comparing the observed accuracy with the expected accuracy within bins of width 0.1. The discrepancy between these two quantities was summarized using the expected calibration error (ECE). The ECE quantifies the difference between expected confidence and the observed accuracy across M bins of predicted probabilities and is defined as:

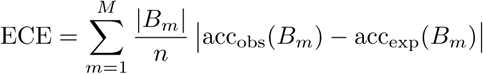

where *B_m_* denotes the set of samples whose predicted confidence scores fall into the *m*-th bin, *|B_m_|* is the number of samples in that bin, and *n* is the total number of samples. Furthermore, the ECE was additionally calculated for the retrospective Nanopore cohort to quantify the discrepancy between the methylation array- and Nanopore-derived predictions.

### 4.7 Classification

Classification was performed using 1,000, 2,500, 5,000, 10,000, 15,000, 20,000, 50,000 and 0.9 x total sequenced CpG sites. For each number of CpG sites the 4 submodels of Tucan were ran 100 times each iteration using a different random subset of CpG sites. The final classification was determined by averaging the different iterations and submodels. For interpretation of the classifications in the retrospective cohort the results at 10,000 CpG sites were used. Classification results were compared to ground truth diagnoses which were provided by the pathologist and based on the combination of histology, immunohistochemistry and molecular analyses.

### 4.8 Model evaluation

Model performance was evaluated using several metrics, including accuracy, precision, recall, and the F_1_ score. Accuracy measures the overall proportion of correctly classified samples (Koo, 2018). Precision and recall were used to assess the model’s specificity and sensitivity, respectively (Koo, 2018). To summarize performance in a single metric that balances precision and recall while accounting for class imbalance, the F_1_ score was computed (Koo, 2018).

Because assessing model performance involved a multiclass classification problem, each class was evaluated using a one-versus-rest strategy, in which the class of interest was treated as the positive class and all remaining classes as negative (Bex, 2021). This yielded per-class performance metrics that were subsequently aggregated using either macro- or micro-averaging (Pahwa, 2017). Macro-averaged metrics assign equal weight to each class, independent of class imbalance, whereas micro-averaged metrics weight classes according to their frequency in the dataset, thereby capturing overall performance across the true class distribution. In this study, macro-averaged, recall, and F_1_ scores were used to ensure equal weighting of all tumor entities irrespective of class frequency, while micro-averaged precision was additionally reported to reflect the overall reliability of positive predictions across the imbalanced cohort.

In addition, we distinguished between global and conditional performance. Global performance was computed across the entire cohort, whereas conditional metrics were calculated within subsets defined by a given confidence threshold *τ*. For each (sub)set, performance was quantified as follows:

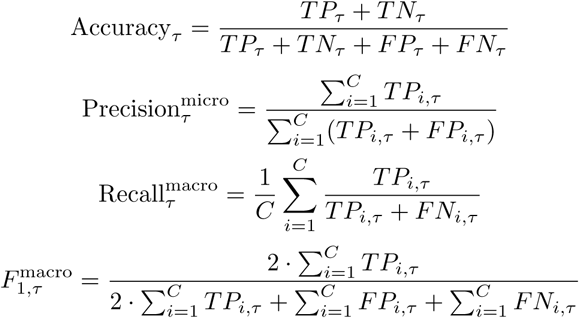

Here, *TP*, *TN*, *FP*, and *FN* denote the true positives, true negatives, false positives, and false negatives, respectively; *i* indicates the class, *C* is the total number of classes, and *τ* denotes the conditional subset based on a confidence threshold.

### 4.9 CNV calling

Sequencing depth was utilized to identify genomic structural variations. Due to the time constraints of same-day sequencing, only shallow coverage could be achieved. Sequencing reads were aligned against the CHM13v2 reference genome using the GATK toolkit, and copy number analysis was performed using GATK-developed CNV calling tools. The genome was divided into non-overlapping bins of 2 megabases, and the sequencing depth was calculated for each bin.

To mitigate technical noise, a panel of normals was constructed from 53 CNV-free Nanopore sequencing samples. Singular value decomposition was applied to this panel to capture the principal sources of systematic variation, and the resulting noise components were subtracted from each sample’s bin-wise depth profile to obtain a denoised signal. Relative coverage for each bin was calculated as the ratio of sample depth to the panel-derived reference depth, yielding a normalized copy ratio signal. These normalized copy ratios were subsequently used for segmentation and structural variant calling.

### 4.10 Sequencing Depth

To determine the minimum sequencing depth required for accurate copy number variation (CNV) detection, raw Nanopore reads were randomly subsampled to generate datasets of increasing depth (3,000; 10,000; 30,000; 100,000; 300,000; and 1,000,000 reads). CNV profiles were computed for each subsampled dataset using the same analytical pipeline applied to the full dataset.

Nanopore derived CNV profiles were evaluated against matched Illumina whole exome sequencing (WES) data. Whole exome libraries were generated starting from 150 ng DNA using the KAPA HyperPrep Kit (Roche) in combination with the KAPA HyperExome capture kit (Roche) and subsequently sequenced on a NovaSeq 6000 sys-tem (2×150 bp, Illumina). The resulting data were processed according to the GATK 4.0 best practices workflow for CNV calling, using a wdl and cromwell based workflow.

The genome was partitioned into 2 Mb bins and concordance was assessed by measuring how closely the Nanopore copy ratios followed the expected identity relationship with WES. Because WES itself contains technical artifacts, and small deviation lack clinical relevance, concordance was defined as falling within a 0.3 band around the identity line *y* = *x*. This threshold captures clinically meaningful CNV deviations while simultaneously avoiding over-penalization from noise inherent to WES. The fraction of bins meeting this criterion was calculated for each sequencing depth.

To evaluate performance beyond random expectation, observed concordance values were compared against a null distribution obtained by random permutation of bin values. This analysis enabled estimation of the sequencing depth at which Nanopore-based CNV calling approached maximal accuracy relative to WES.

### 4.11 DNA Extraction

Total DNA was isolated using the AllPrep DNA/RNA/Protein Mini Kit (Qiagen) on a Qiacube (Qiagen) according to manufacturers instructions. Since native DNA sequencing of FFPE derived DNA is not possible using the oxford Nanopore technologies platform, only DNA derived from fresh frozen material was used. Genomic DNA (*>* 500 ng) isolated pediatric solid tumor tissue was processed for Nanopore sequencing on the GridION/PromethION P2i/PromethION P24 using the Oxford Nanopore Native Barcoding Kit (V14). High–molecular weight gDNA underwent FFPE-repair, end-prep, and bead-based clean-up, followed by ligation of barcodes and adapter ligation according to the manufacturers instructions. Barcoded libraries were pooled (up to 6 samples per minION R10.4.1 or up to 24 samples per R10.4.1 flow cell) and loaded after standard priming with Flow Cell Flush reagents. Sequencing was performed with Dorado high-accuracy basecalling, with 5mC/5hmC detection, followed by alignment to the CHM13v2.0 reference. For each sample, approximately 200 million basecalled bases (corresponding to 0.07x genome coverage 25,000 relevant CpG sites) were generated to enable classification by Tucan. Sequencing performance and barcode assignments were monitored in MinKNOW.

## 5 Consent for publication

The research was approved by the Biobank and Data access committee (BDAC, PMCLAB2023.0437) of the Princess Máxima Center for pediatric oncology. All included patients provided written informed consent for participation in the biobank (International Clinical Trials Registry Platform: NL7744; https://onderzoekmetmensen.nl/en/trial/21619).

## 6 Patient identity

Patient identifiers for samples obtained from the Princess Máxima Center biobank were already pseudonymized prior to access. The link between the provided identifiers and patient identities is not accessible outside the originating biobank. For publicly available datasets, the original identifiers provided in the repository metadata were retained.

## 7 Data availability

Pediatric patient data from the Princess Máxima are currently being pre-pared for submission to the European Genome–Phenome Archive (EGA) and will be made available under accession number xxxxxxx. Public data used for training are available at GEO, TARGET and TCGA under accession numbers: GSE131677, GSE140686, GSE161692, GSE186487, GSE192967, GSE196228, GSE214568, GSE37362, GSE78732, MTAB 9875, TARGET-CCSK, TARGET-NBL, TARGET-OS, TARGET-RT, TARGET-WT, TCGA-ACC, TCGA-KICH, TCGA-KIRC, TCGA-KIRP, TCGA-PCPG, TCGA-SKCM, TCGA-TGCT, TCGA-THCA.

## 8 Code availability

Source code of the Tucan prediction tool as a Python package can be found at: https://github.com/UMCUGenetics/tucan, together with the link to download the trained model used in this work.

## 9 Supplementary information

Most supplementary information is included within the manuscript itself (see Section 14). Supplementary Table S2 presents a collapsed version of the sample overview table. The complete, uncollapsed table is provided separately as an Excel file.

## 10 Acknowledgements

We acknowledge the EpSSG MyKids Study for providing DNA samples for cases PMC4, PMC53, PMC54, and PMC58. We thank the Princess Máxima Center Biobanking facility and the Big Data Core for the storage and provision of samples and data. This project was funded by a project grant from Stichting Kinderen Kanker Vrij (KiKa).

## 11 Conflict of interest

Jeroen de Ridder is co-founder of Cyclomics BV, a genomics company. Jeroen de Ridder, Carlo Vermeulen and Marc Pagés-Gallego are named inventors on a patent application related to the Sturgeon classifier.

## 12 Author Contribution

Conceptualization: M.J., B.T., J.d.R. and L.K., Methodology: M.J., M.P., and C.V., Software: M.J., M.P., and L.K., Validation: M.J., and L.K., Formal analysis: M.J., and L.K., Investigation: M.J., M.v.T., and E.d.R., Resources: L.H., U.F., R.d.K., M.S., P.K. R.v.E., H.M., M.v.N., Data curation: M.J., L.H., U.F., R.d.K., M.S., P.K. R.v.E., H.M., M.v.N. and L.K., Writing - Original draft: M.J. and L.K., Writing - Review & Editing: All authors., Visualization: M.J., Supervision: C.V., B.T., J.d.R. and L.K., Project administration: L.K., Funding acquisition: B.T., J.d.R. and L.K.

## 14 Supplementary Information

**Table S1:**
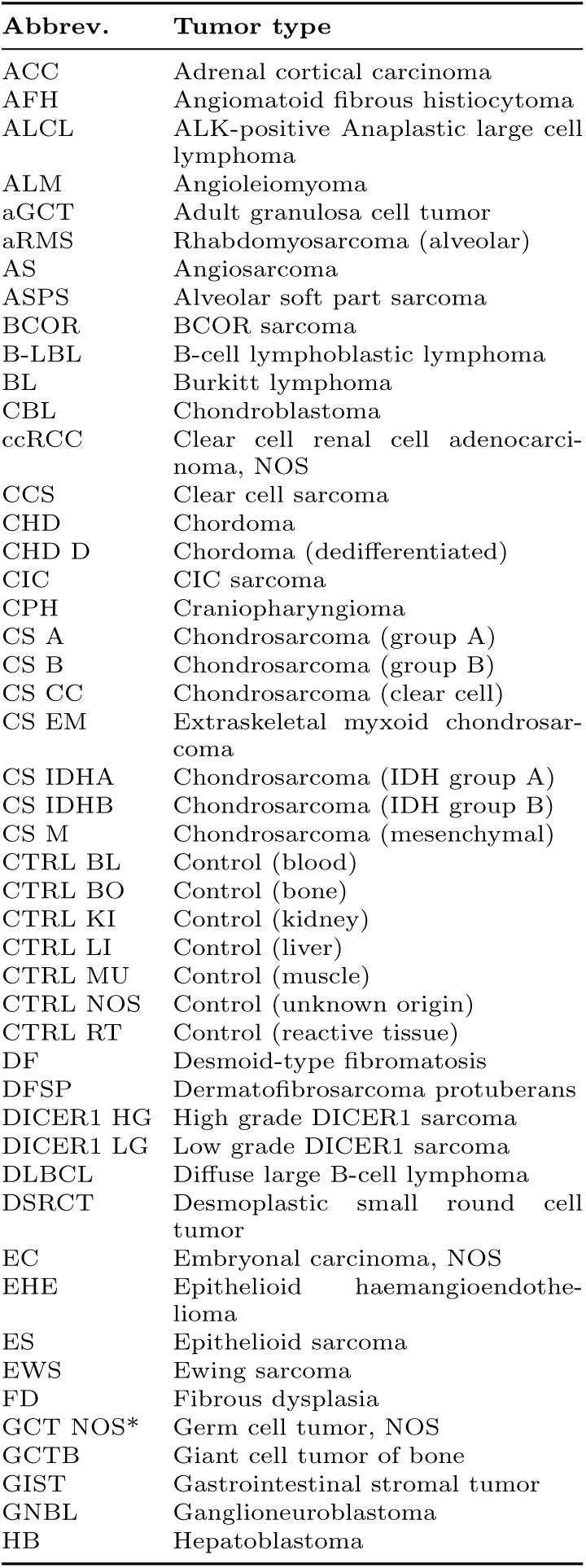
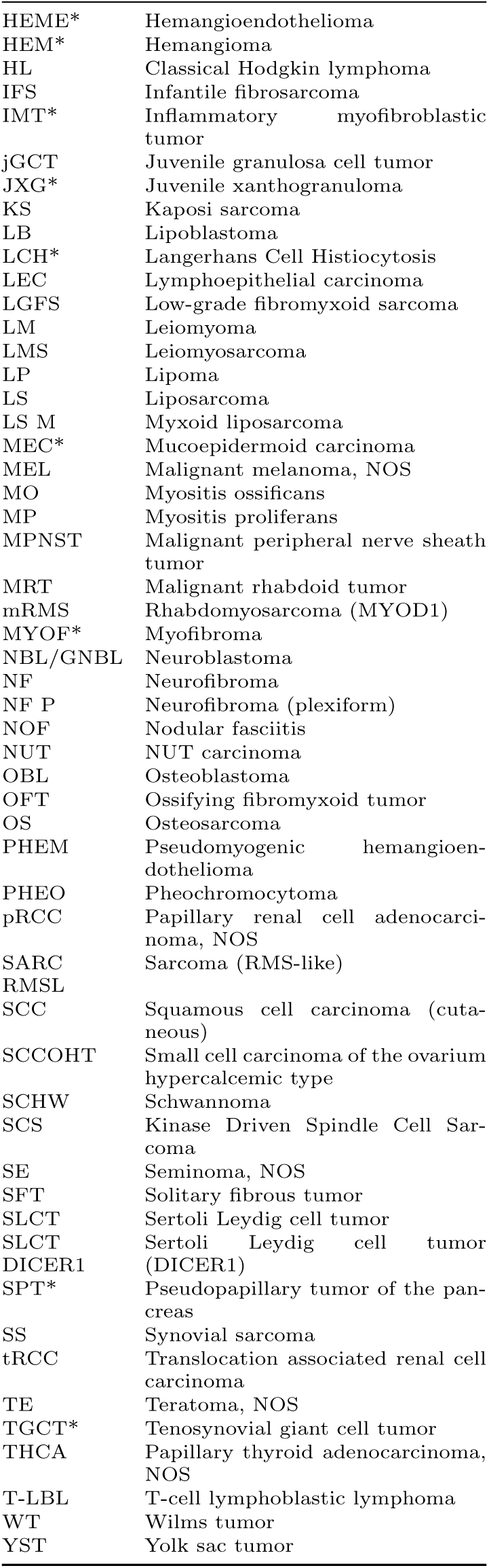
Tumor types and corresponding abbreviations used in Tucan and the manuscript. Tumor entities are listed alphabetically by abbreviation. Subtypes included in the manuscript but not implemented in the Tucan classifier are indicated with an asterisk (*).

**Fig. S1:**
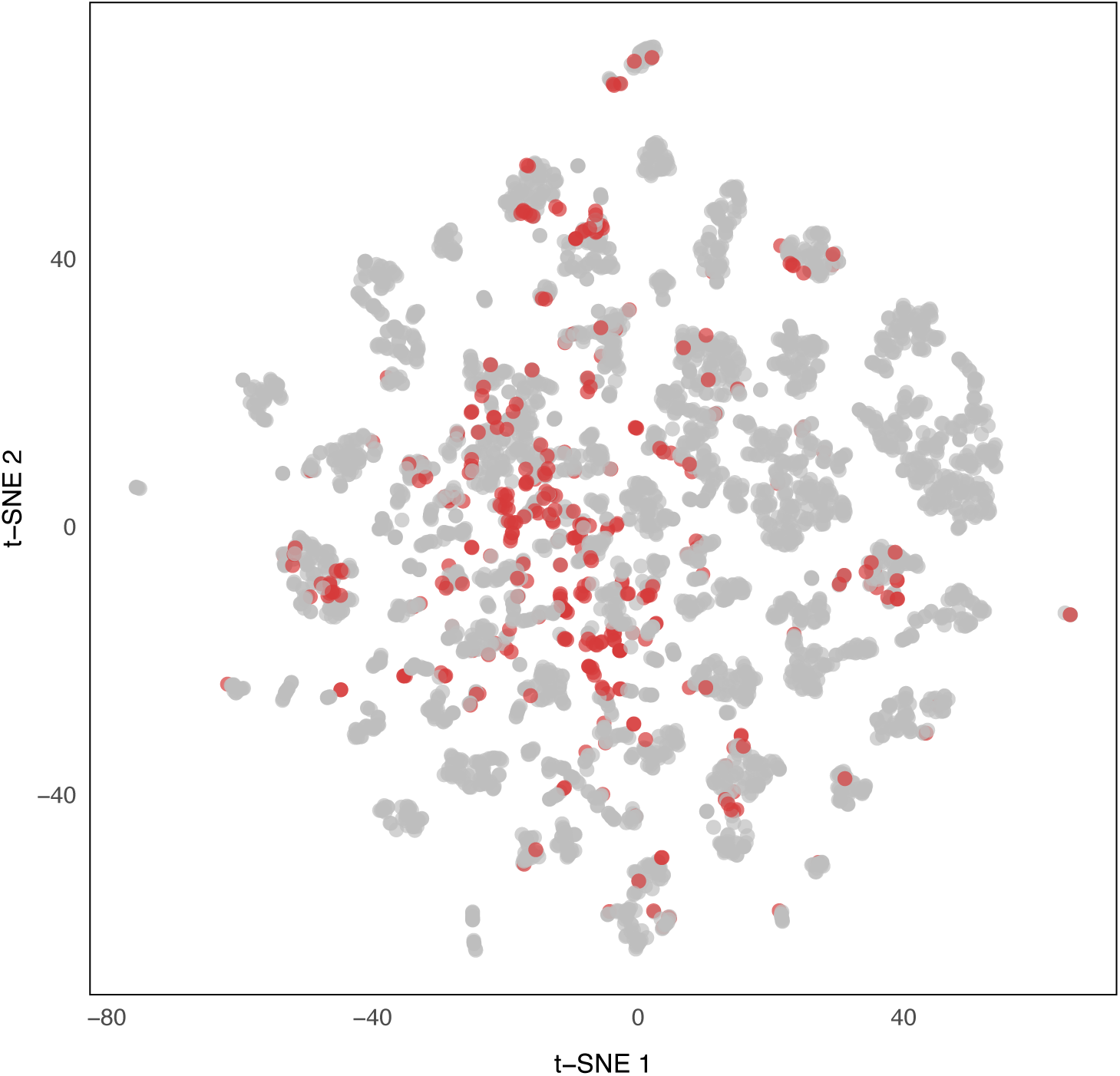
Training cohort structure prior to data curation. t-SNE visualization of the methylation array training cohort based on the 5,000 most variable CpG sites. Each point represents a sample in the training dataset. Samples shown in grey were retained for model training, whereas samples highlighted in red were removed during the subsequent data filtering procedure.

**Fig. S2:**
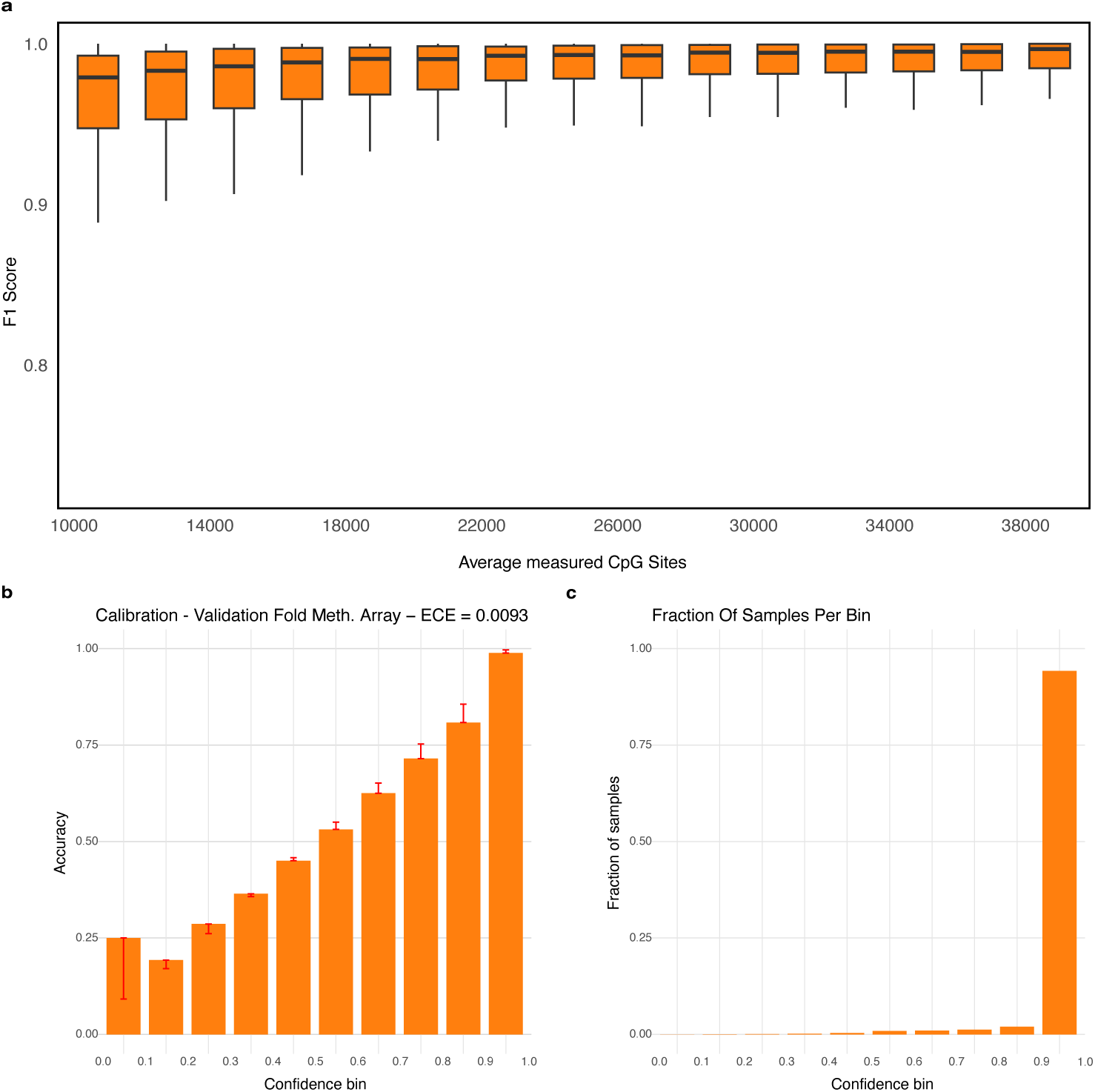
Performance and Bin Statistics - Methylation Array Validation Data. **(a)** F1 score as a function of the number of measured CpG sites (sequencing depth), computed on simulated sparse representations of patient samples. Performance improves with increasing CpG coverage. **(b)** Calibration plot with expected calibration error (ECE) for the validation fold of the methylation array data. The bars show observed accuracies within each bin, with red segments indicating the corresponding calibration error per bin. **(c)** The fraction of samples that contributed to calculating the conditional performance per bin.

**Fig. S3:**
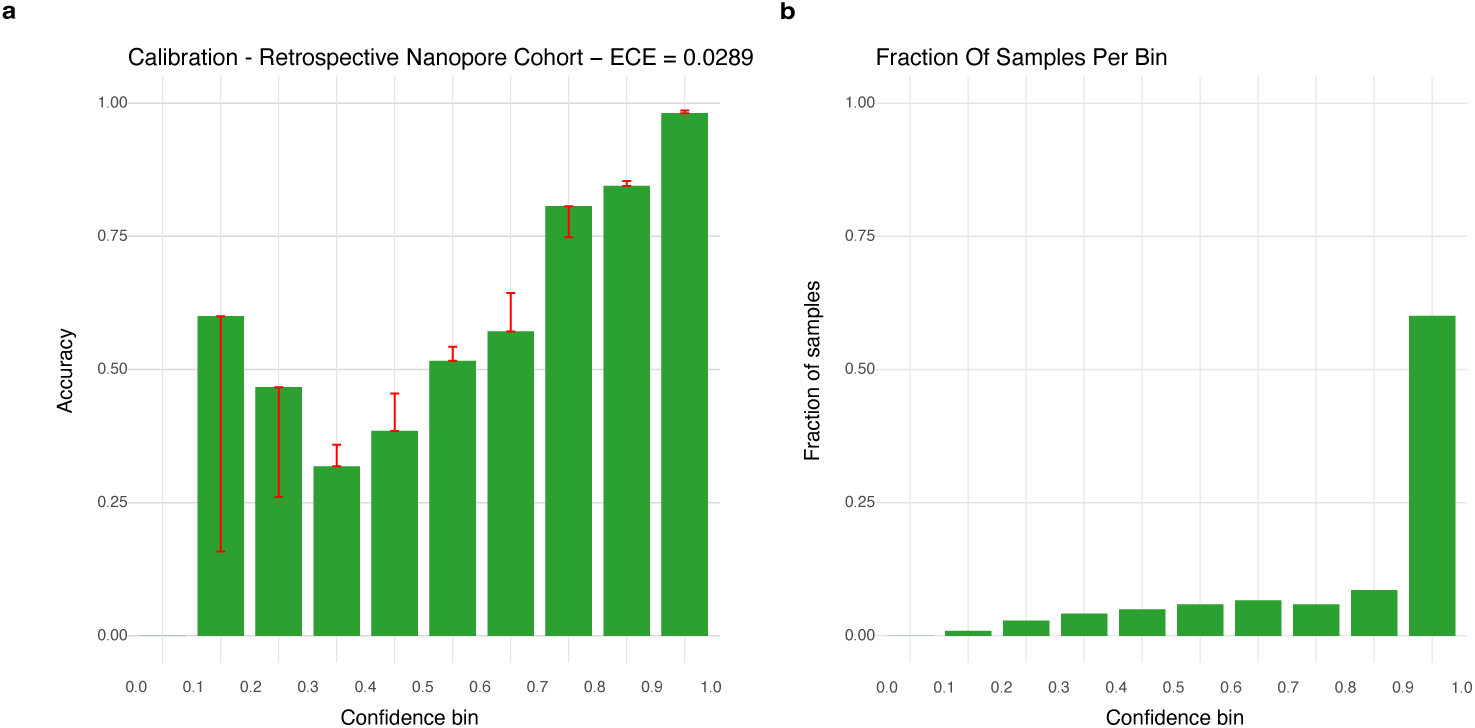
Bin Statistics - Retrospective Nanopore Samples. **(a)** Calibration plot with expected calibration error (ECE) for the retrospective Nanopore samples. The bars show observed accuracies within each bin, with red segments indicating the corresponding calibration error per bin. **(b)** The fraction of samples that contributed to calculating the conditional performance per bin.

**Fig. S4:**
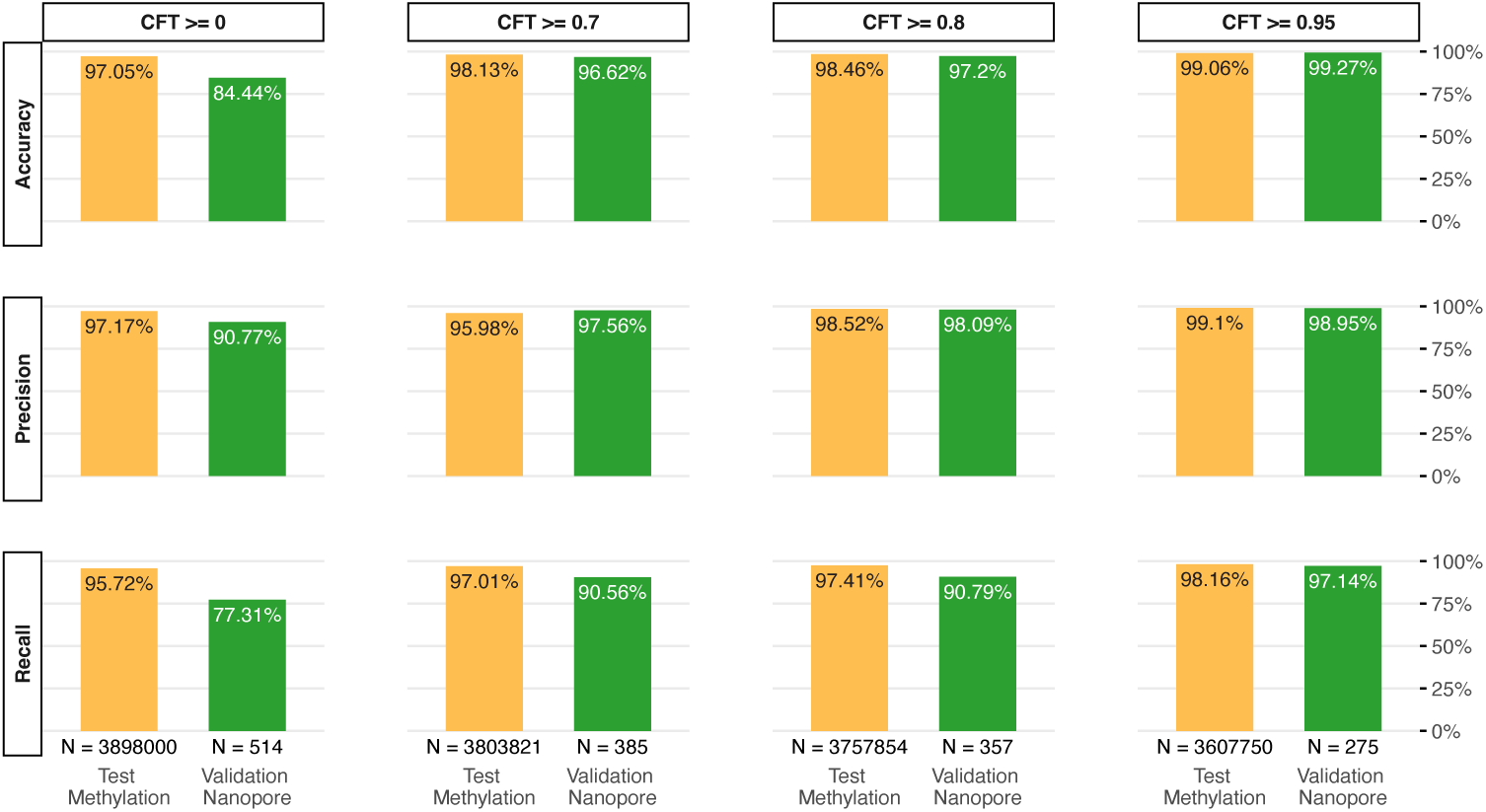
Performance of confidence-thresholded predictions. Accuracy, precision, and recall are shown for the test methylation array data (yellow) and an independent retrospective Nanopore cohort (green) at increasing confidence thresholds (CFT *≥* 0, 0.7, 0.8, and 0.95). For each threshold, metrics are computed conditionally, considering only samples with prediction confidence greater than or equal to the specified threshold. For both datasets, predictions are derived from simulated sparse representations of patient samples, generated by subsampling CpG sites. For the methylation array (yellow), metrics and sample counts (N) are computed at the level of individual sparse samples, whereas for the Nanopore cohort (green), sparse predictions are aggregated back to their corresponding original patient samples prior to metric calculation.

**Fig. S5:**
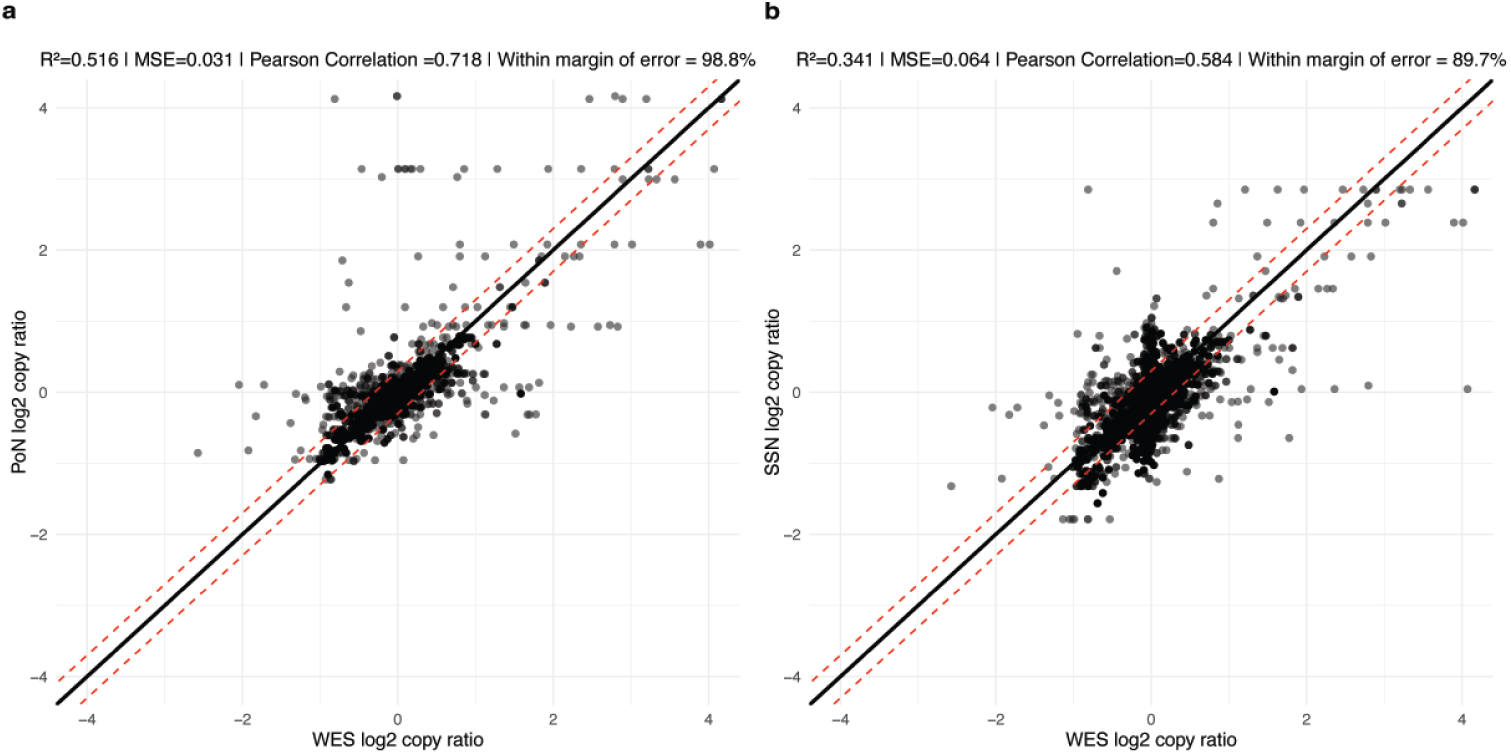
Normalization based on panel of normals outperforms deeply single sequenced sample normalization in CNV detection. Correlation of copy ratios of called CNV segments (binned at 2 Mb) between Whole Exome Sequencing (WES) and **(a)** the Panel of Normals (PoN), and **(b)** the deeply single-sequenced sample (SSN). Each dot represents the copy ratio of a called segment within a 2 Mb bin. The solid black line indicates the line of identity (*y* = *x*), while the red dashed lines represent the predefined margin of error (*y* = *x ±* 0.3), within which differences are considered clinically equivalent. Summary statistics are shown in each panel (*R*^2^, MSE, and Pearson correlation). Analyses are based on 38 samples with available WES, PoN, and SSN data.

**Fig. S6:**
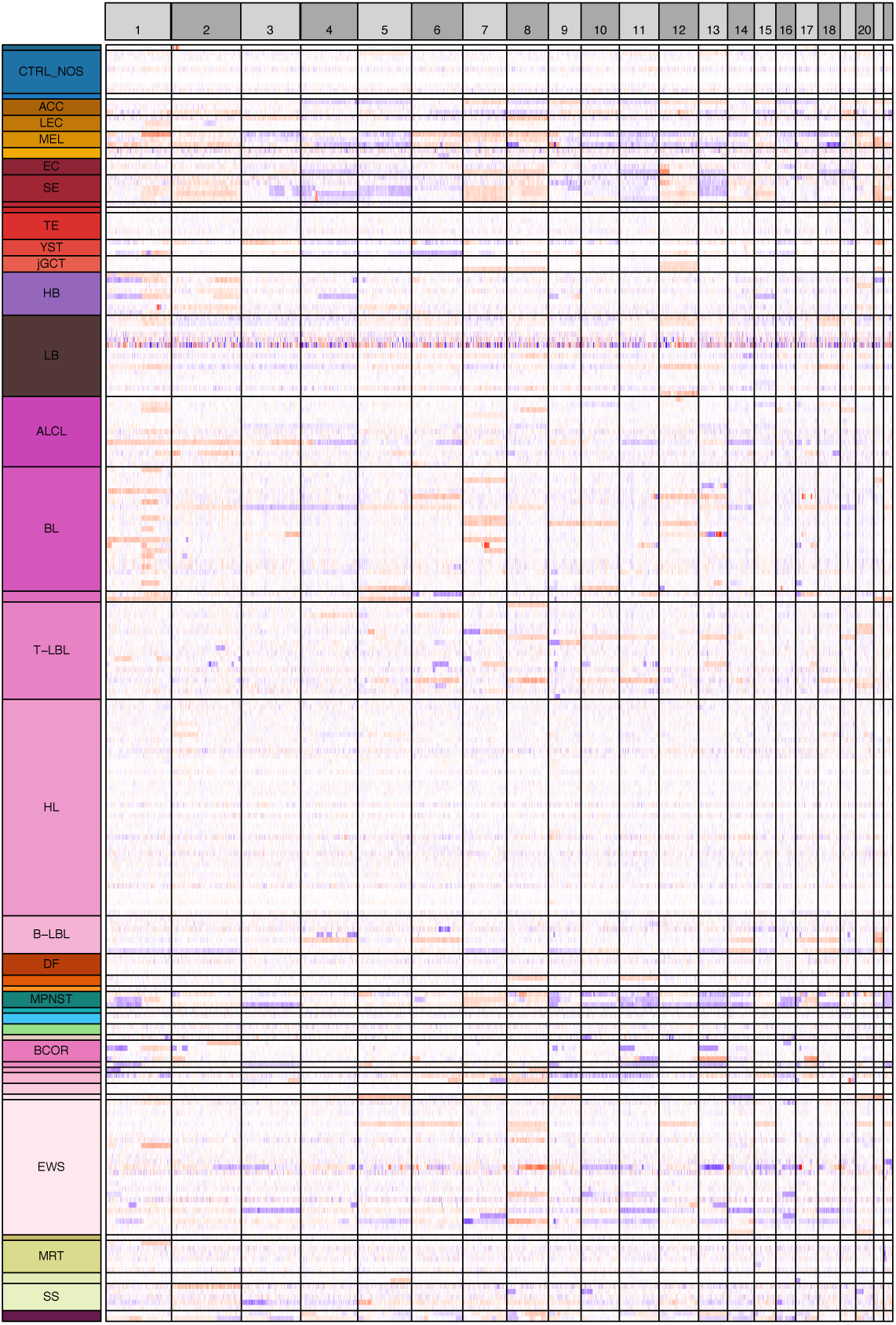
Heatmaps of copy number variations (CNVs) per subtype per chromosome. Each row represents an individual sample, color-coded by subtype. Columns correspond to chromosomes, with alternating light and dark grey shading for visual clarity. Red indicates copy number gains, blue indicates copy number losses, and white denotes neutral regions.

**Table S2:** Collapsed overview of training samples and tumor subtype annotations. This table presents a collapsed overview of samples used for model training, including sample identifiers, study of origin, tumor subtype abbreviations, and an indicator of whether each sample was included in the training set. For readability, only a summarized version is shown here. The complete table containing all samples and annotations is provided as a separate Excel file (Supplementary Table S2).

| Sample Name | Identifier | Study | Subtype Abbreviation | In Training |
| --- | --- | --- | --- | --- |
| 100994770004_R02C02 | source_1 | MTAB.9875 | ABC | TRUE |
| 100994770004_R04C02 | source_4 | MTAB.9875 | GCTB | TRUE |
| 100994770004_R05C02 | source_6 | MTAB.9875 | GCTB | FALSE |
| 100994770004_R06C01 | source_7 | MTAB.9875 | GCTB | TRUE |
| 100994770005_R03C02 | source_11 | MTAB.9875 | ABC | TRUE |
| 100994770005_R05C01 | source_12 | MTAB.9875 | OBL | TRUE |
| 101103430066_R01C01 | source_16 | MTAB.9875 | GCTB | FALSE |
| 101103430066_R02C01 | source_18 | MTAB.9875 | ABC | TRUE |
| 101103430066_R03C01 | source_20 | MTAB.9875 | OBL | TRUE |
| 101103430066_R03C02 | source_21 | MTAB.9875 | OBL | TRUE |
| 101103430066_R04C01 | source_22 | MTAB.9875 | OS | FALSE |
| 101103430066_R04C02 | source_23 | MTAB.9875 | OBL | TRUE |
| 101103430066_R05C02 | source_25 | MTAB.9875 | OBL | TRUE |
| 101103430084_R06C02 | source_27 | MTAB.9875 | ABC | TRUE |
| 101103430084_R03C01 | source_30 | MTAB.9875 | GCTB | TRUE |
| 101103430084_R03C02 | source_31 | MTAB.9875 | OBL | TRUE |
| 101103430084_R04C01 | source_32 | MTAB.9875 | ABC | TRUE |
| 101103430084_R05C01 | source_34 | MTAB.9875 | ABC | TRUE |
| 101103430084_R06C01 | source_36 | MTAB.9875 | OBL | TRUE |
| 101103430087_R01C01 | source_37 | MTAB.9875 | GCTB | TRUE |

**Fig. S7:**
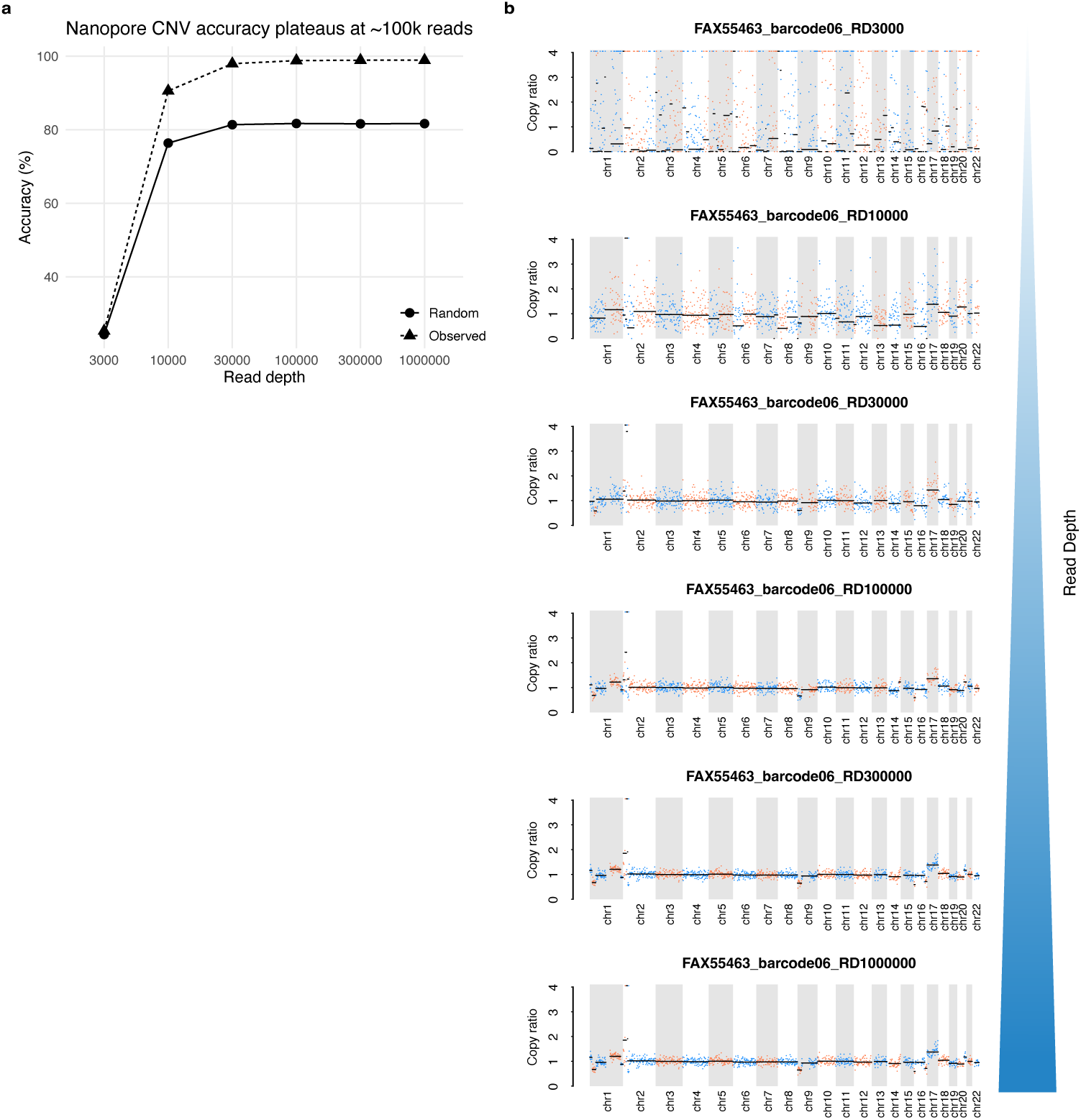
Nanopore CNV accuracy over sequencing depth. **(a)** The dotted line shows the accuracy of CNV detection from Nanopore sequencing at increasing read depths, benchmarked against WES as reference (agreement within ±0.3 copy ratio). Accuracy is shown for the observed copy-ratio profiles as well as for a random control, generated by permuting bin-wise copy-ratio values across genomic positions to disrupt positional concordance while preserving the global distribution of copy ratios. Because the majority of genomic bins are copy-neutral, the random baseline exhibits non-zero accuracy. Accuracy of the observed profiles increases with sequencing depth and stabilizes at approximately 100,000 reads, reaching 99%. **(b)** CNV plots for a neuroblastoma sample with MYCN amplification on chromosome 2 across different read depths. The horizontal axis denotes chromosomes (alternating grey and white blocks), and the vertical axis shows the copy ratio. Dots represent copy ratios calculated in 2 Mbp bins, with black horizontal lines indicating segmentation boundaries. Colors alternate between blue and red per called segment for visual clarity. MYCN amplification is readily detectable, and the CNV profile stabilizes at approximately 100k reads.

**Fig. S8:**
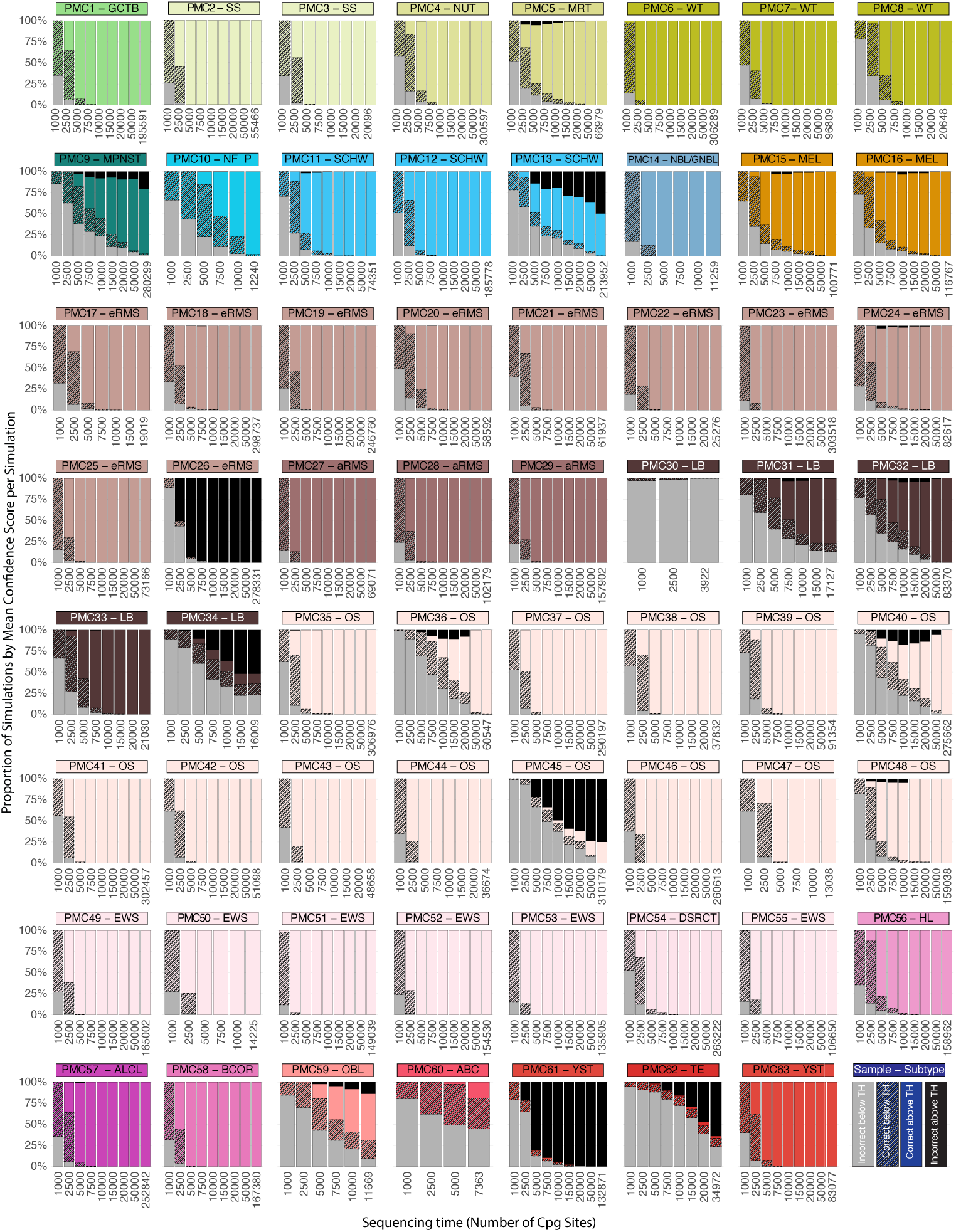
Tucan classification of simulated prospective samples across sequencing depth at a confidence threshold of 0.7. For each sample, repeated subsampling generated multiple simulations at increasing sequencing depths. Each simulation was classified and assigned a mean confidence score, which was compared to the 0.7 threshold. The horizontal axis represents sequencing depth (number of CpG sites), and the vertical axis represents the proportion of simulations per sample falling into each classification category at the respective depth. Samples are color-coded by subtype. Solid subtype colors indicate correct classifications above the confidence threshold, striped subtype colors indicate correct classifications below the threshold, grey denotes incorrect classifications below the threshold, and black indicates incorrect classifications above the threshold.

**Fig. S9:**
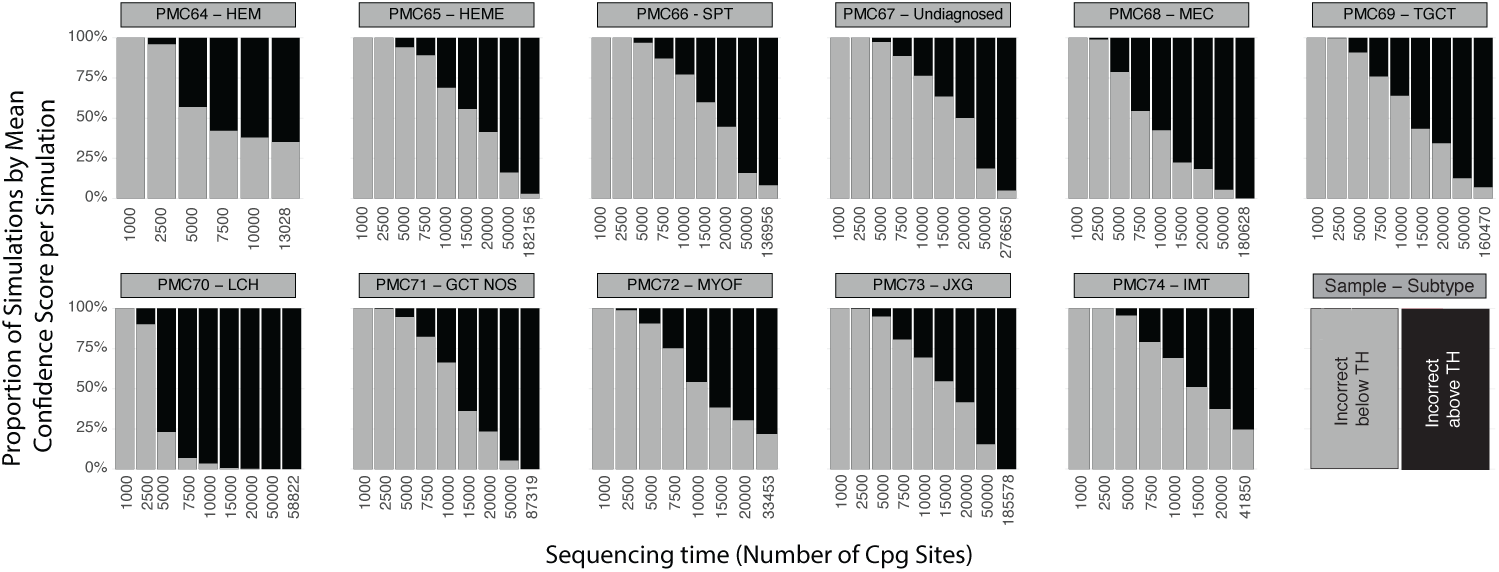
Tucan classification of simulated prospective samples not included in classifier training across sequencing depth at a confidence threshold of 0.7. For each sample, repeated subsampling generated multiple simulations at increasing sequencing depths. Each simulation was classified and assigned a mean confidence score, which was compared to the 0.7 threshold. The horizontal axis represents sequencing depth (number of CpG sites), and the vertical axis represents the proportion of simulations per sample falling into each classification category at the respective depth. As these subtypes were not part of the training set, only incorrect classifications are observed: grey denotes incorrect classifications below the confidence threshold, and black indicates incorrect classifications above the threshold.

**Fig. S10:**
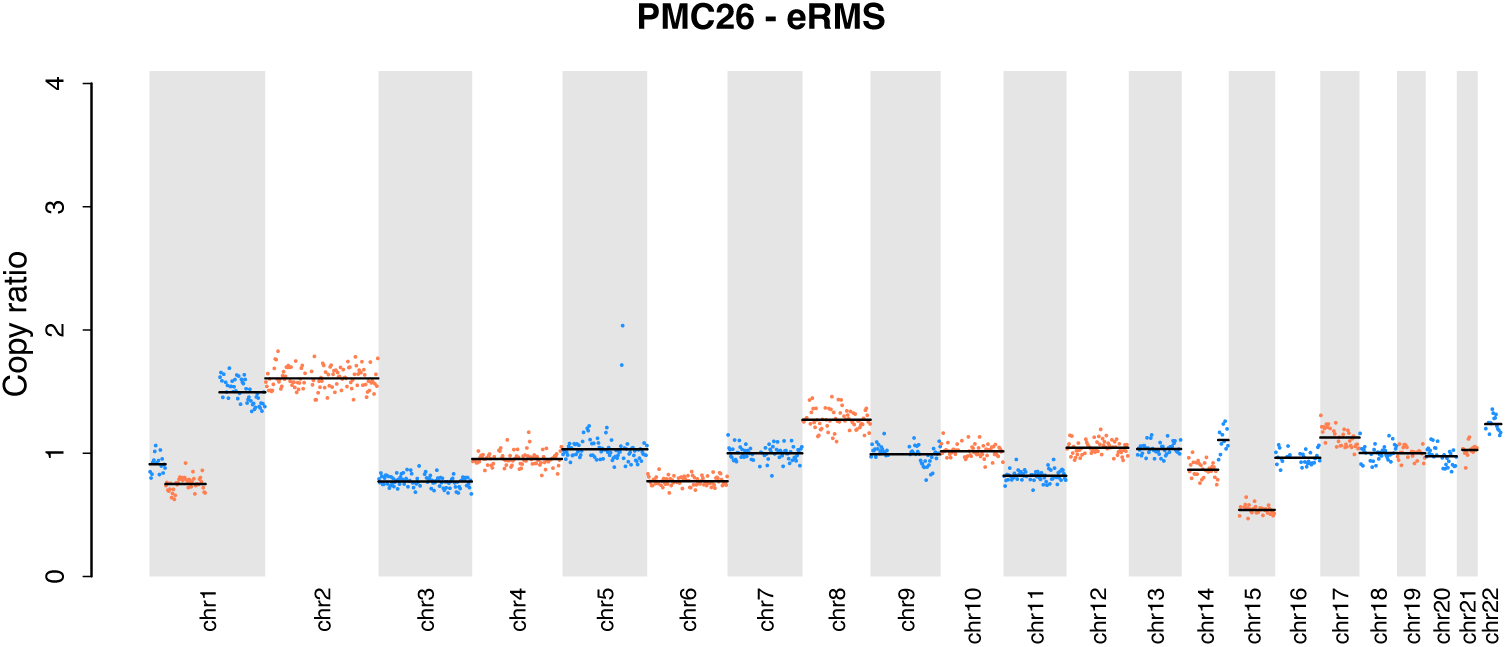
CNV profile of prospective patient PMC26. The horizontal axis denotes chromosomes (alternating grey and white blocks), and the vertical axis shows the normalized copy ratio. Dots represent copy-ratio values calculated in 2 Mbp genomic bins, with black horizontal lines indicating segmentation boundaries. Dots alternate between blue and red according to the assigned segment for visual clarity. The profile demonstrates large-scale chromosomal gains and losses, consistent with the characteristic copy-number pattern of the embryonal rhabdomyosarcoma (eRMS) subtype.

**Fig. S11:**
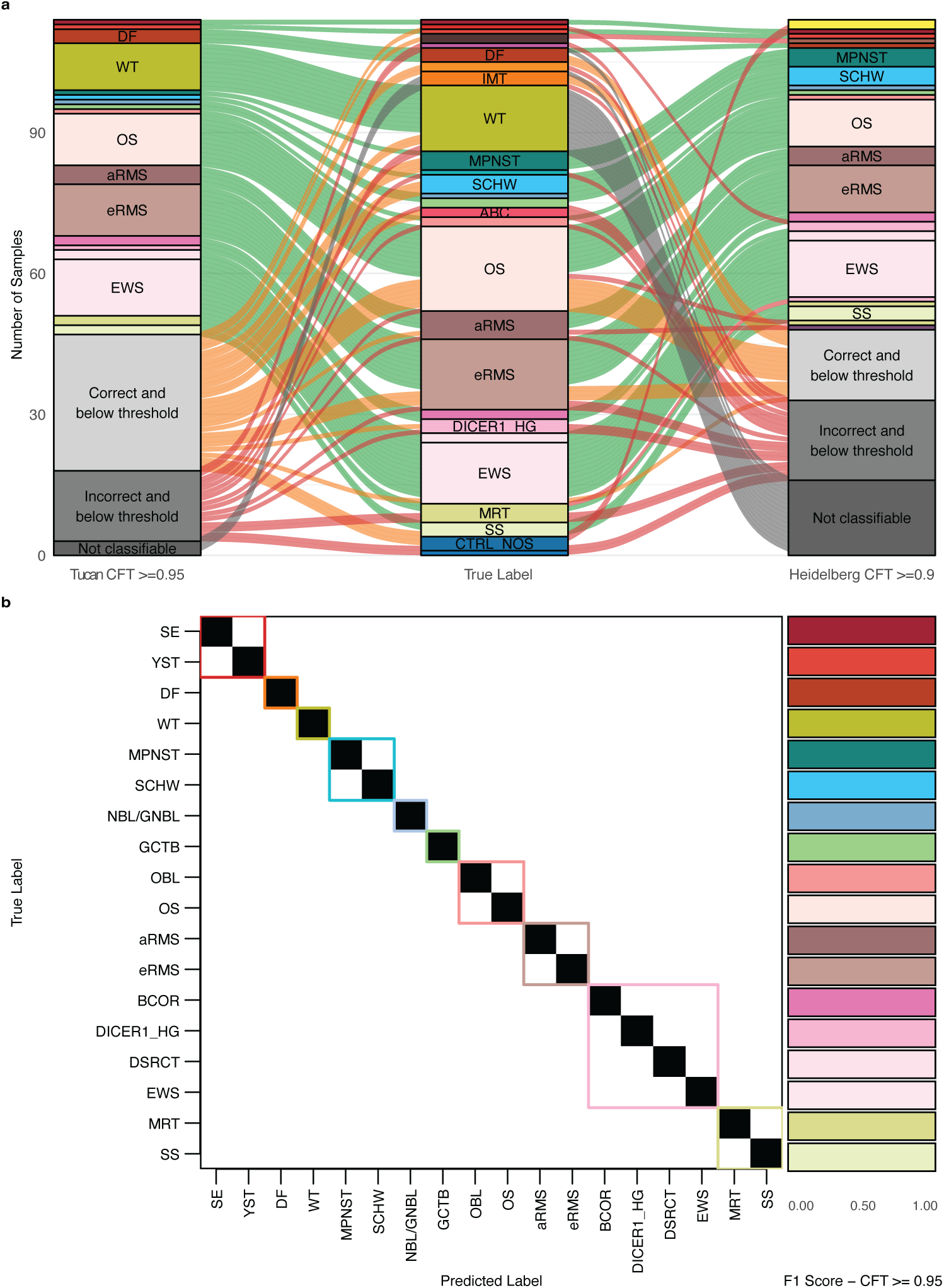
Performance of Tucan and Heidelberg Classifier - Retrospective Methylation Array Samples. **a** Alluvial plot comparing Tucan predictions at a confidence threshold (CFTs) *≥* 0.95 (left) with Heidelberg predictions at a CFT *≥* 0.9 (right). True tumor labels are shown in the center column. Predictions are colored by subtype, with low-confidence predictions and non-classifiable samples shown in grey. Flow colors indicate prediction accuracy: green, correct (*≥* CFT); orange, correct (*<* CFT); red, incorrect; and grey, not classifiable. **b** Correlation matrix of Tucan’s predictions evaluated at a CFT *≥* 0.95. Subtypes are color-coded, with true labels displayed along the vertical axis and predicted labels along the horizontal axis. The corresponding F1-scores for each subtype are displayed to the right of the matrix.

## Notes

### Competing Interest Statement

Jeroen de Ridder is co-founder of Cyclomics BV, a genomics company. Jeroen de Ridder, Carlo Vermeulen and Marc Pages-Gallego are named inventors on a patent application related to the Sturgeon classifier.

### Author Declarations

The research was approved by the Biobank and Data access committee (BDAC, PMCLAB2023.0437) of the Princess Maxima Center for pediatric oncology. The biobank collection is registered as a clinical trial in the Netherlands and has been approved by the Dutch Medical Ethical Commission. All included patients provided written informed consent for participation in the biobank (International Clinical Trials Registry Platform: NL7744; https://onderzoekmetmensen.nl/en/trial/21619).

